# Healthy dynamics of CD4 T cells may drive HIV resurgence in perinatally-infected infants on antiretroviral therapy

**DOI:** 10.1101/2022.02.09.22270686

**Authors:** Sinead E. Morris, Renate Strehlau, Stephanie Shiau, Elaine J. Abrams, Caroline T. Tiemessen, Louise Kuhn, Andrew J. Yates, the EPIICAL Consortium and the LEOPARD study team

## Abstract

In 2019 there were 490,000 children under five living with HIV. Understanding the dynamics of HIV suppression and rebound in this age group is crucial to optimizing treatment strategies and increasing the likelihood of infants achieving and sustaining viral suppression. Here we studied data from a cohort of 122 perinatally-infected infants who initiated antiretroviral treatment (ART) early after birth and were followed for up to four years. These data included longitudinal measurements of viral load (VL) and CD4 T cell numbers, together with information regarding treatment adherence. We previously showed that the dynamics of HIV decline in 53 of these infants who suppressed VL within one year were similar to those in adults. However, in extending our analysis to all 122 infants, we find that a deterministic model of HIV infection in adults cannot explain the full diversity in infant trajectories. We therefore adapt this model to include imperfect ART adherence and natural CD4 T cell decline and reconstitution processes in infants. We find that individual variation in both processes must be included to obtain the best fits. We also find that, perhaps paradoxically, infants with faster rates of CD4 reconstitution on ART were more likely to experience resurgences in VL. Overall, our findings highlight the importance of combining mathematical modeling with clinical data to disentangle the role of natural immune processes and viral dynamics during HIV infection.

**Author Summary:** For infants infected with HIV at or near birth, early and continued treatment with antiretroviral therapy (ART) can lead to sustained suppression of virus and a healthy immune system. However many treated infants experience viral rebound and associated depletion of CD4 T cells. Mathematical models can successfully capture the dynamics of HIV infection in treated adults, but many of the assumptions encoded in these models do not apply early in life. Here we study data from a cohort of HIV-positive infants exhibiting diverse trajectories in response to ART. We show that wide-ranging outcomes can be explained by a modified, but still remarkably simple, model that includes both the natural dynamics of their developing immune systems and variation in treatment adherence. Strikingly, we show that infants with strong rates of recovery of CD4 T cells while on ART may be most at risk of virus resurgence.

## Introduction

In 2019 there were 490,000 children under five living with HIV, and 150,000 newly diagnosed cases [1]. Although infants receiving antiretroviral treatment (ART) can suppress viral load (VL), eventually the cessation of treatment leads to HIV rebound, due to reactivation of latently-infected cells. Nevertheless, early initiation of ART can lead to extended periods of suppression in the absence of treatment – for example, over 22 months in the case of the ‘Mississippi Child’ and 8.75 years in a South African participant of the Children with HIV Early antiRetroviral therapy (CHER) trial [2, 3]. Therefore, understanding the dynamics of HIV suppression and rebound following ART initiation in young infants is crucial for optimizing treatment strategies and increasing the likelihood of achieving and sustaining viral suppression.

We previously showed that a simple biphasic model of VL decay captures the early dynamics of HIV decline in perinatally-infected infants on ART and that these dynamics are similar to those in adults [4]. However, models applied to dynamics of infection in adults over longer timescales typically encode assumptions that do not extend to infants [5–12]. First, CD4 T cell dynamics in adults are typically described as a balance between a constant total rate of influx and a constant *per capita* rate of loss, leading to steady trajectories in the absence of infection. In contrast, HIV-uninfected infants experience a natural, exponential decline in CD4 T cell numbers per unit volume of blood as the immune system matures [13]. Second, perinatally-infected infants undergo a transient period of CD4 T cell reconstitution upon ART initiation, during which numbers quickly recover to those of HIV-uninfected infants [14]. This short-lived process cannot be captured by the constant CD4 recruitment term exploited in many models of adult infection. Third, the standard assumption that ART is completely effective in blocking new infection of cells may not hold true for young infants, due to challenges in treatment adherence. Thus, canonical models of HIV suppression and rebound in adults must be modified for infants to include potential reductions in ART efficacy, and more complex dynamics of CD4 T cell numbers.

Here we model the dynamics of HIV infection in a cohort of perinatally-infected infants from Johannesburg, South Africa who initiated ART early in life. We extend a simple deterministic model of HIV suppression and rebound in adults to incorporate incomplete treatment adherence and dynamics of natural CD4 T cell decline and reconstitution. By fitting this model to longitudinal viral RNA and CD4 T cell data, we estimate rates of reactivation and reconstitution. We also show that individual variation in CD4 reconstitution rates are an important factor driving variation in HIV suppression and resurgence characteristics across infants, in addition to ART adherence. Overall, our results demonstrate the complex interplay between natural immune processes and HIV dynamics, and highlight the importance of mathematical modeling in disentangling these factors.

## Methods

### Data

The LEOPARD study has been described previously [4, 15]. Briefly, 122 perinatally-infected infants were enrolled at the Rahima Moosa Mother and Child Hospital in Johannesburg, South Africa, between 2014 and 2017. The majority began ART within two weeks of birth (median age: 2.5 days; interquartile range (IQR): 1–8), and were followed for up to four years. VL (HIV RNA copies ml^-1^) and CD4 T cell concentrations (cells *µ*l^-1^) in the blood were sampled over time, and various clinical covariates were also recorded, including the infant’s pre-treatment CD4 percentage, the mother’s VL and CD4 count after delivery, and the mother’s prenatal ART history (full list in Table S1).

With these data, we previously identified a subset of 53 infants who successfully suppressed VL within one year [4, 16], with suppression defined as having at least one VL measurement below the 20 copies ml^-1^ detection threshold of the RNA assay. Here, we are interested in the interplay between natural CD4 T cell dynamics and infection processes, and whether and how this interplay determines whether an infant achieves suppression and/or experiences VL rebound. We have therefore broadened our analysis to all 122 infants.

In addition to the data described previously, we include information relating to ART adherence that was obtained at each study visit (details in SI). For each drug in each infant’s ART regimen – with the recommended, and most common, being zidovudine (AZT), lamivudine (3TC), and (i) nevirapine (NVP) in the first four weeks of treatment, then (ii) ritonavir-boosted lopinavir (LPV/r) after four weeks – we estimated a percentage adherence by comparing the weight of medicine returned to the expected amount returned assuming perfect adherence. Less than 100% adherence can result from missed doses or ‘under-dosing’ (giving too little medicine at each dose), whereas greater than 100% adherence can occur through over-dosing or problems with drug tolerance (infants may spit up bad-tasting medicine, therefore requiring repeat dosing). For many visits, adherence could not be calculated because leftover medicine was not returned. The adherence estimates are therefore influenced by many unobserved factors and, given uncertainty in how the quantitative estimates map to actual adherence, we instead defined a categorical variable that labeled adherence estimates greater than 90% as ‘good’, and estimates less than 90% as ‘poor’. Using 85% and 95% as alternative thresholds for good adherence did not alter our findings. With this course-grained approach, some missing values could be manually labelled based on physician commentary from accompanying questionnaires (for example, if substantial gaps in dosing were noted, adherence was labeled as poor). We then summarized the average adherence of each infant as the most frequently reported category (good or poor) across their time series.

### Ethics statement

All protocols for the LEOPARD study were approved by the Institutional Review Boards of the University of the Witwatersrand and Columbia University. Written informed consent was obtained from mothers for their own and their infants’ participation.

### Model

We describe HIV dynamics in an infant on ART using a deterministic ordinary differential equation (ODE) model (Fig 1A) [7, 12]. We assume that CD4 target cells, *T* (*t*) (measured as the concentration of cells per *µ*l of blood, but from here on referred to as ‘cells’ or ‘counts’ for brevity), undergo background growth and loss according to a function *θ*(*t, T*), and are infected by free virus, *V* (*t*), at *per capita* transmission rate *β* (Table 1). During ART this transmission is blocked with efficacy *ϵ*_1_, where *ϵ*_1_ *<* 1 reflects incomplete adherence of reverse transcriptase inhibitor drugs that block infection of new cells (for example, NVP and AZT). A proportion (1 - *ρ*) of the newly infected target cells seed the latent reservoir; the remaining fraction, *ρ*, either become productively infected or die through abortive infection [17]. Cells from this heterogeneous infected population, *I*(*t*), are lost at an average rate *d*, and produce virus at an average rate (1 − *ϵ*_2_)*p*, where *p* subsumes the fraction of cells that are productively infected and *ϵ*_2_ *<* 1 reflects any failure of protease inhibitors (for example, LPV/r) to block the production of infectious virus. The infected cell population is also boosted by reactivation of the latent reservoir, at rate *a*. We do not explicitly model the number of latently infected cells due to uncertainties in the rates of proliferation and loss in this population, and a lack of available data to estimate these parameters. Finally, free virus is lost at rate *c*. This system can be represented by the following equations

**Table 1:**
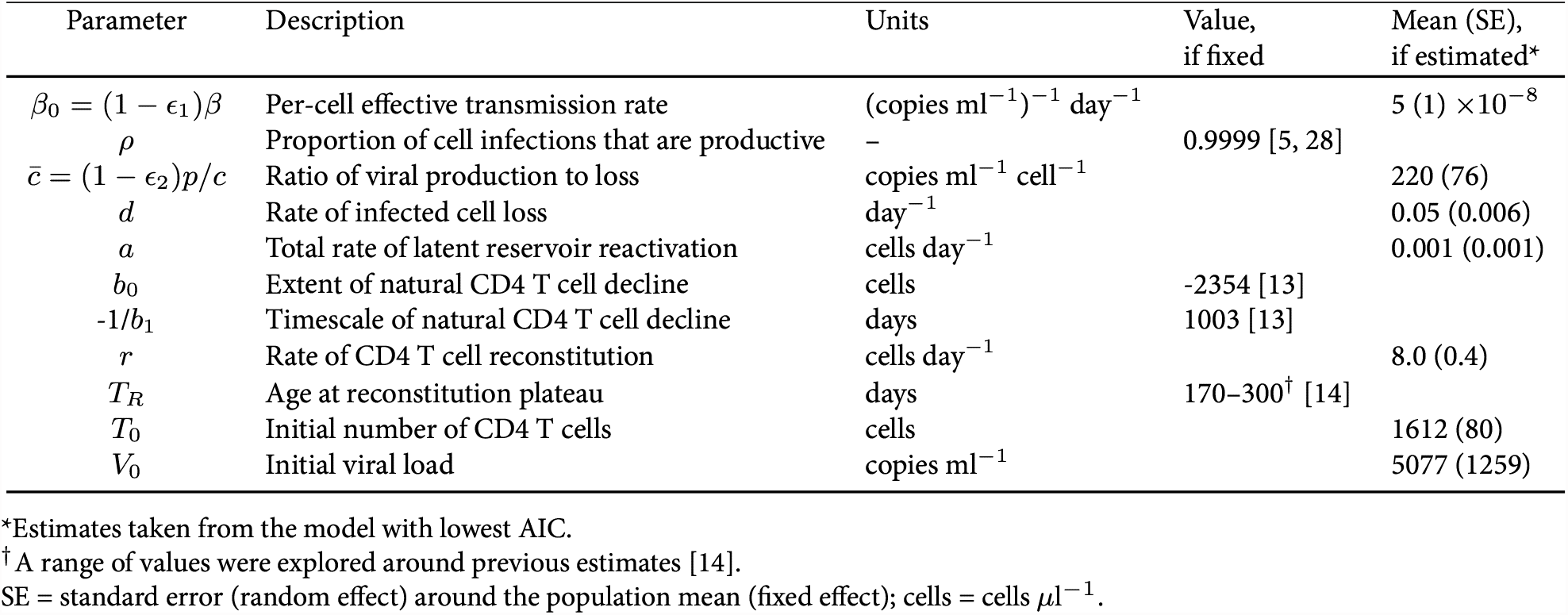
Model parameters and population-level estimates.

**Figure 1:**
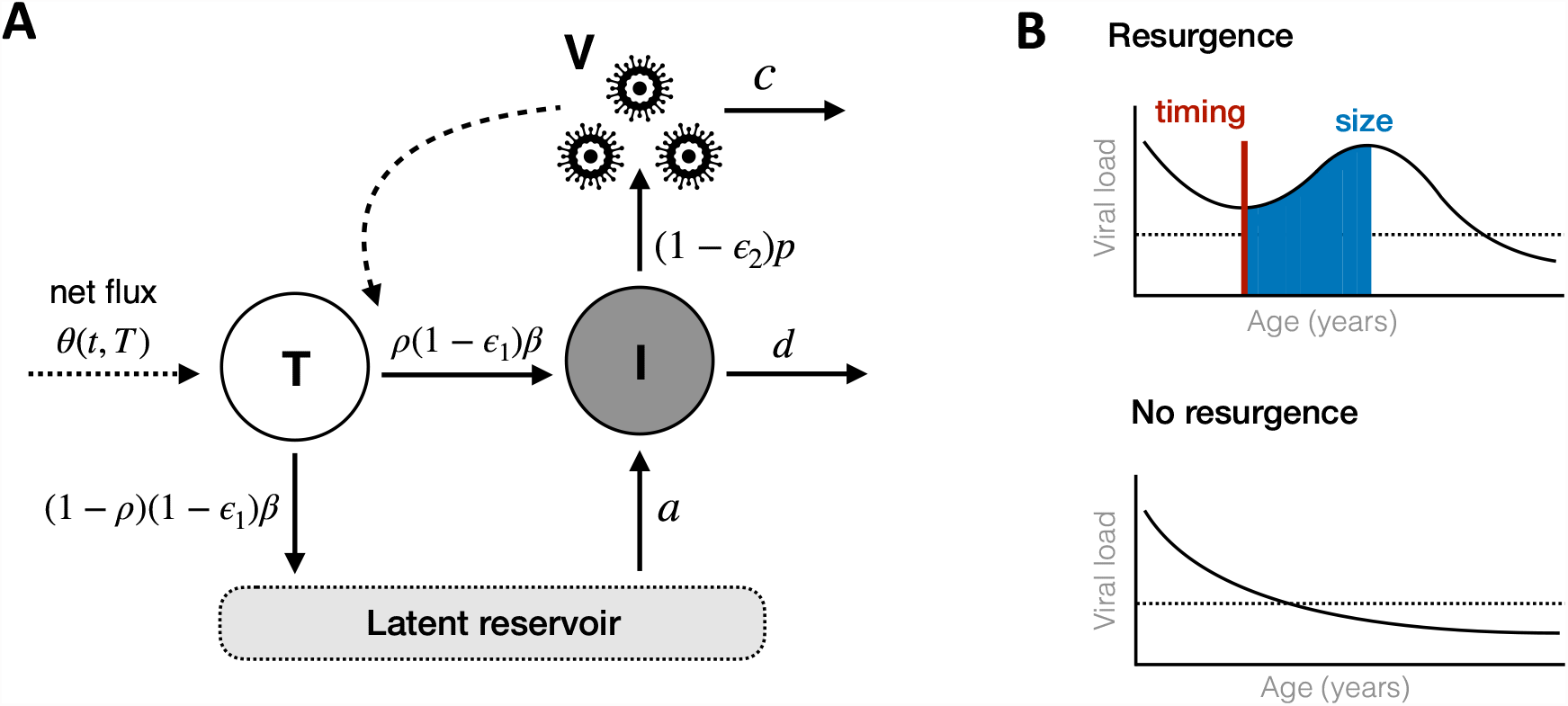
Model framework and analysis schematic. (A) Framework for the infection model with rate constants. CD4 target cells (T) are infected by free virus (V) and either become productively infected cells (I) or latently infected cells. Productively infected cells produce free virus whereas latently infected cells do not; but latently infected cells can become reactivated at a later point to join the productively infected cell population. CD4 target cells also undergo reconstitution and natural decline processes according to *θ*(*t, T*). Further details are given in the text. (B) Schematic illustrating the definition of viral resurgence (top) compared to no resurgence (bottom). The timing of resurgence is defined as the time at which VL first starts increasing (vertical red line), and the size of resurgence is the total integrated VL during the upslope period (blue shaded region). The dashed horizontal line represents the assay detection threshold.

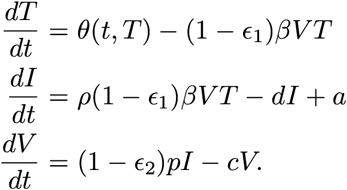

The reproduction number for this model at ART initiation is

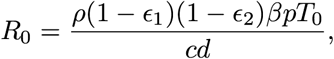

where *T*_0_ is the initial CD4 count (assuming contribution from the latent reservoir is negligible at this time).

In the simplest case we assume all rate parameters are constant over time. However, we also investigate an extension of this model that incorporates a delay in reactivation of the latent reservoir,

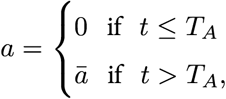

where *T*_*A*_ is the time to reactivation in days. Assuming the rate of virus turnover is faster than that of CD4 T cells [18], we reduce the model to the following system

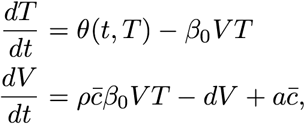

where *β*_0_ = (1 − *ϵ*_1_)*β* and 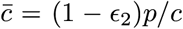 (derivation in the SI). The compound parameter 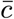 represents the contribution of each infected cell to the total viral load, through its rate of production of virions and the average time they persist in circulation. Both *β*_0_ and 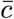 incorporate ART efficacy (through *ϵ*_1_ and *ϵ*_2_), and thus infant ART adherence.

Finally, we extend the model for young infants through the term governing background CD4 T cell growth and loss, *θ*(*t, T*). In models of infection dynamics in adults, *θ*(*t, T*) typically takes the form *λ*−*d*_*T*_ *T*, where *λ* and *d*_*T*_ are constant rates representing cell influx and natural decay processes, respectively. These forms lead to steady trajectories in the absence of infection. For infants, we propose an alternative *θ*(*t, T*) that instead accounts for the exponentially declining concentration of CD4 T cells that is observed as HIV-uninfected infants age [13], and (ii) the transient recovery in CD4 counts experienced by HIV-infected infants during the early stages of ART [14]. First, the natural decline in CD4 T cells can be captured by an exponential decay function

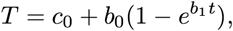

where *c*_0_, *b*_0_ and *b*_1_ are constant parameters that have been independently estimated in a cohort of 80 uninfected children in Germany, including 39 aged between 2 months and 4 years [13]. This function also captured CD4 T cell dynamics in 381 South African children, of whom 300 were aged between 2 weeks and 5 years [19]. Second, the additional reconstitution of the CD4 T cell pool in HIV-infected infants can be modeled as a transient increase in cell counts during the early stages of ART, i.e.

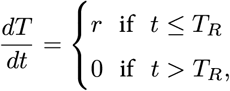

where *r* is the constant rate of reconstitution and *T*_*R*_ is the time to reach healthy levels [14]. Combining these processes of reconstitution and the natural decline of CD4 T cell counts in infants gives 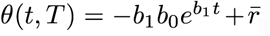, and

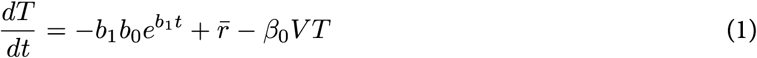

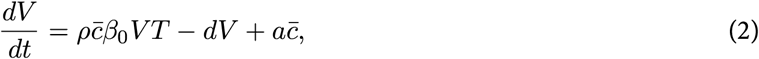

where

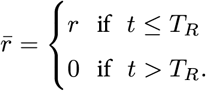

### Model fitting and comparisons

We fit equations 1 and 2 to the VL and CD4 T cell data from all 122 infants using a nonlinear mixed effects approach. All VL observations below the detection threshold were treated as censored values, and we assumed both *V* (*t*) and *T* (*t*) were lognormally distributed [20, 21]. Given the relative infrequency of CD4 T cell measurements, we fixed four parameters across all individuals (Table 1): three that governed the reconstitution and natural dynamics of target cells (*T*_*R*_, *b*_0_ and *b*_1_), and the proportion of newly infected cells that become productively infected (*ρ*). All other parameters were estimated and allowed to have both fixed and random effects. In subsequent analyses we estimated fixed and random effects for *T*_*R*_. We also examined the importance of individual variation in adherence and CD4 T cell recovery by comparing the best fit model to three alternative models in which *β*_0_, 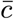 or *r* were fixed across all infants.

Following exploratory fits, each estimated parameter was assumed to follow a lognormal distribution, with the exception of *a* which followed a logit-normal distribution with pre-specified upper bound, and *r* and *T*_*R*_ which followed normal distributions. We verified that the random effects for all estimated parameters were normally distributed, using the Shapiro-Wilk test. Guided by the exploratory fits, we allowed *β*_0_ and *d* to be correlated, and assumed all other parameters were independent. We confirmed the identifiability of all parameters [22], and conducted additional sensitivity analyses by varying each chosen parameter in turn and re-simulating the model, while keeping all other parameters fixed. We used these simulations to assess the sensitivity of model predictions to our choice of fixed parameters. Model fitting and parameter estimation were implemented in Monolix 2020R1 [21]. Downstream analyses and plotting were conducted in R version 4.03 [23], with the `deSolve, cowplot, patchwork` and `tidyverse packages` [24–27]. All details needed to reproduce our analyses are given in the SI.

We compared the statistical support for different models using the Akaike Information Criterion (AIC). For model *i*, AIC_*i*_ = 2*k* − 2 ln *L*, where *k* is the number of estimated parameters, ln *L* is the maximum loglikelihood, and lower AIC values indicate stronger statistical support. We assessed the relative support for model *i* using ΔAIC_*i*_ = AIC_*i*_ - AIC_*min*_, where AIC_*min*_ is the minimum AIC value across all models. Differences greater than five indicate substantially greater support for the model with AIC_*i*_ = AIC_*min*_. For the favored model, we used the individual-specific parameter estimates to predict VL and CD4 T cell trajectories for each child. These trajectories extended either to the end of our study period or two years after their last observation, whichever was earlier. We then compared how viral infection and the natural decline in CD4 T cells mediated the overall VL and CD4 T cell dynamics. We calculated the relative contributions of new viral infection and natural decline to decreases in CD4 T cell numbers as

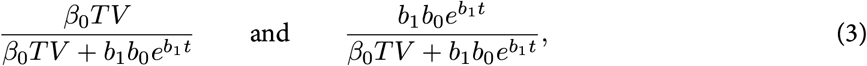

respectively. Similarly, the relative contributions of new viral infection and latent reservoir reactivation to increases in the number of productively infected cells were

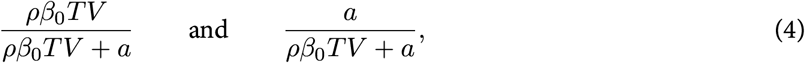

respectively.

### Statistical analyses

We tested for statistical associations between model parameters, clinical covariates (Table S1), and the risk of VL resurgence – defined as any predicted increase in VL following initiation of ART (Fig 1B). We chose VL resurgence as our indicator of imperfect viral control rather than VL rebound (any predicted increase in VL following initial suppression of HIV) due to the small number of infants experiencing the latter (5/122 infants experienced rebound compared to 52/122 experiencing resurgence). We defined the timing of VL resurgence as the first point at which the model predicted an increase in VL, and the size of resurgence as the total integrated VL during the upslope period (Fig 1B). We then tested for associations using Spearman correlations between pairs of quantitative variables, the Kruskal Wallis test between quantitative and categorical variables, and Chi-squared tests between pairs of categorical variables. We adjusted for multiple testing using the Benjamini-Hochberg correction.

## Results

The model for adult infection, with *θ*(*t, T*) = *λ* − *d*_*T*_ *T*, was a poor fit to the infant data, particularly the CD4 T cell counts (Fig S1, Table 2). We therefore used the model adapted for infant infection, with 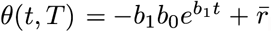, in all further analyses. First, we verified that the infant model with fixed time to reconstitution plateau, *T*_*R*_, and constant rate of latent reactivation, *a*, was structurally identifiable (see Table 1 and SI) [22]. We initially fixed *T*_*R*_ = 225 days across all infants, following previous modeling of CD4 reconstitution in another cohort of HIV-infected infants who initiated ART 82 days after birth, on average [14]. We refitted the model with different fixed values and verified that the best fits were obtained when *T*_*R*_ = 225 days (ΔAIC_*i*_ = 8.3 relative to the next best model with *T*_*R*_ = 223 days; Table S2). Including random effects for *T*_*R*_ improved model fits, although also estimating the fixed effect (i.e. estimating the population average of *T*_*R*_) did not (Table 2). This is likely due to the increased complexity introduced by estimating an additional parameter. Similarly, including a delay in reactivation of the latent reservoir did not improve model fits. We therefore focus on the model with a constant rate of reactivation from the latent reservoir and random effects for *T*_*R*_ around a predefined population mean of 225 days. With this model, VL predictions were marginally sensitive to all fixed parameters (*T*_*R*_, *b*_0_, *b*_1_ and *ρ*; Fig S2), whereas CD4 T cell dynamics were sensitive to those governing healthy T cell reconstitution and decline (*T*_*R*_, *b*_0_ and *b*_1_) but robust to changes in the proportion of new infections that become productively infected cells (*ρ*; Fig S3).

**Table 2:** Model comparisons. AIC values (ΔAIC) are quoted relative to the minimum AIC value across all models. The model with ΔAIC = 0 is the model with lowest AIC and thus has most statistical support. See Table 1 for parameter definitions.

| Model* | $\Delta\text{AIC}$ |
| --- | --- |
| Constant $a$; fixed $T_R = 225$ days with random effect | 0.0 |
| Constant $a$; fixed $T_R = 225$ days without random effect | 7.1 |
| Constant $a$; estimated $T_R$ | 27.1 |
| Time-varying $a$, fixed $T_R = 225$ days without random effect | 31.5 |
| $\theta(t, T) = \lambda - d_T T$ | 365.4 |

\*Unless stated otherwise, $\theta(t, T) = -b_1 b_0 e^{b_1 t} + \bar{r}$ .

Strikingly, our relatively simple deterministic model captured the wide variation in infant VL trajectories, including monotonic decreases to suppression, eventual suppression following transient increases in VL, and brief periods of suppression with a subsequent rebound in VL (Fig 2). Later, or multiple, rebound occurrences were generally not so well captured. These behaviors may be due to repeated fluctuations in treatment adherence or stochastic processes driving delayed reactivation of the latent reservoir, neither of which are included in the model. Initially, new infections were the major contributor to growth of the productively infected cell population (Eqn 4; Fig S4). However, in almost all infants the importance of new infection events was eventually superseded by reactivation from the latent reservoir, although this displacement was delayed by viral resurgence events and prolonged CD4 T cell recovery (Fig S5).

**Figure 2:**
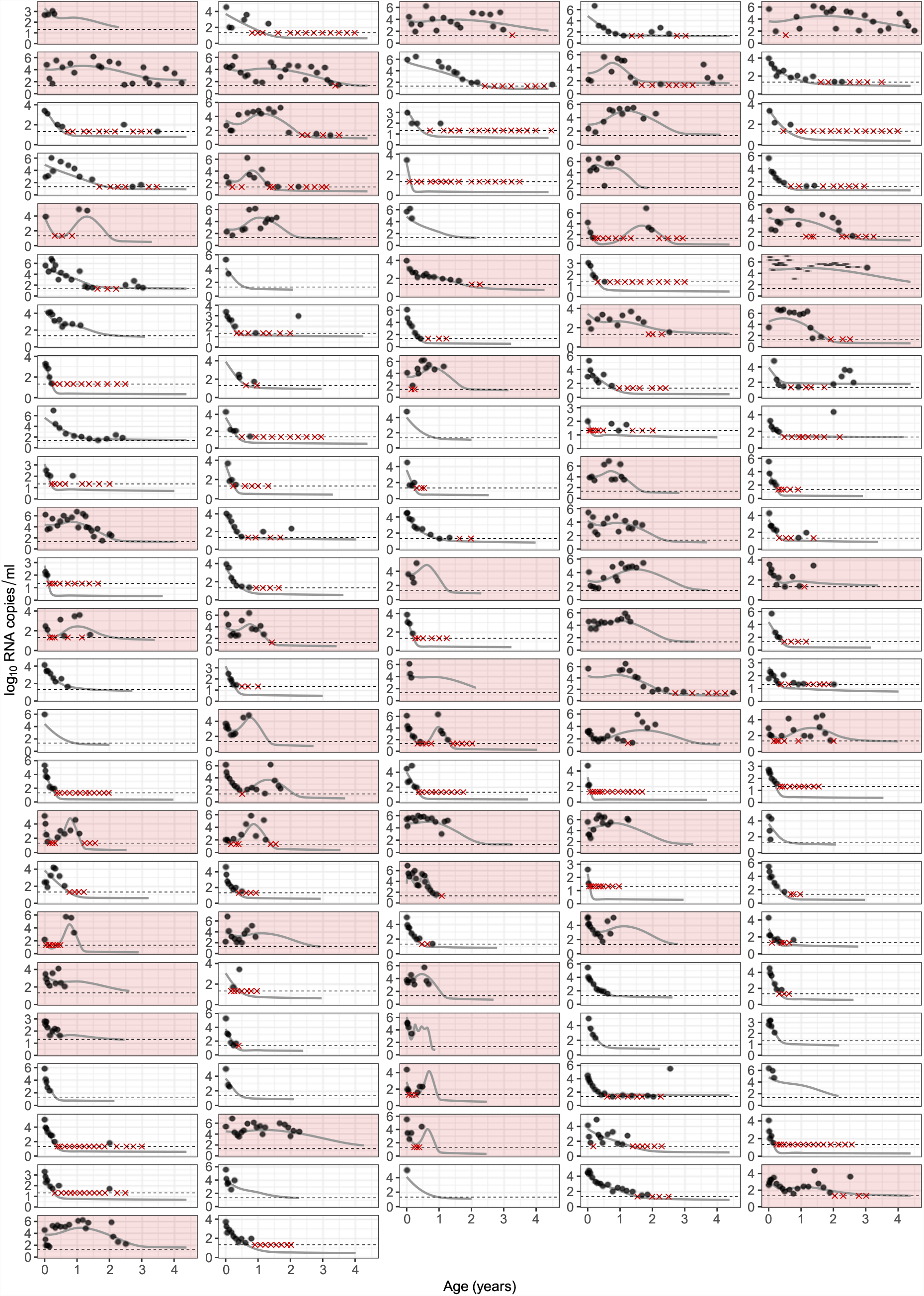
Model fits for RNA observations. Each panel represents a different infant; points represent the data; and solid lines are the model fits. The dashed horizontal line is the detection threshold of the RNA assay, and red crosses are censored observations below this threshold. Panels shaded in red are infants who experienced viral resurgence (i.e. at least one period of increasing VL).

The majority of infants experienced a transient increase in CD4 T cell counts followed by a steady decline; these patterns were well captured by the model (Fig 3). The decline in CD4 T cells was almost always driven by natural processes, although the contribution of new infections increased during periods of VL resurgence (Eqn 3; Fig S6).

**Figure 3:**
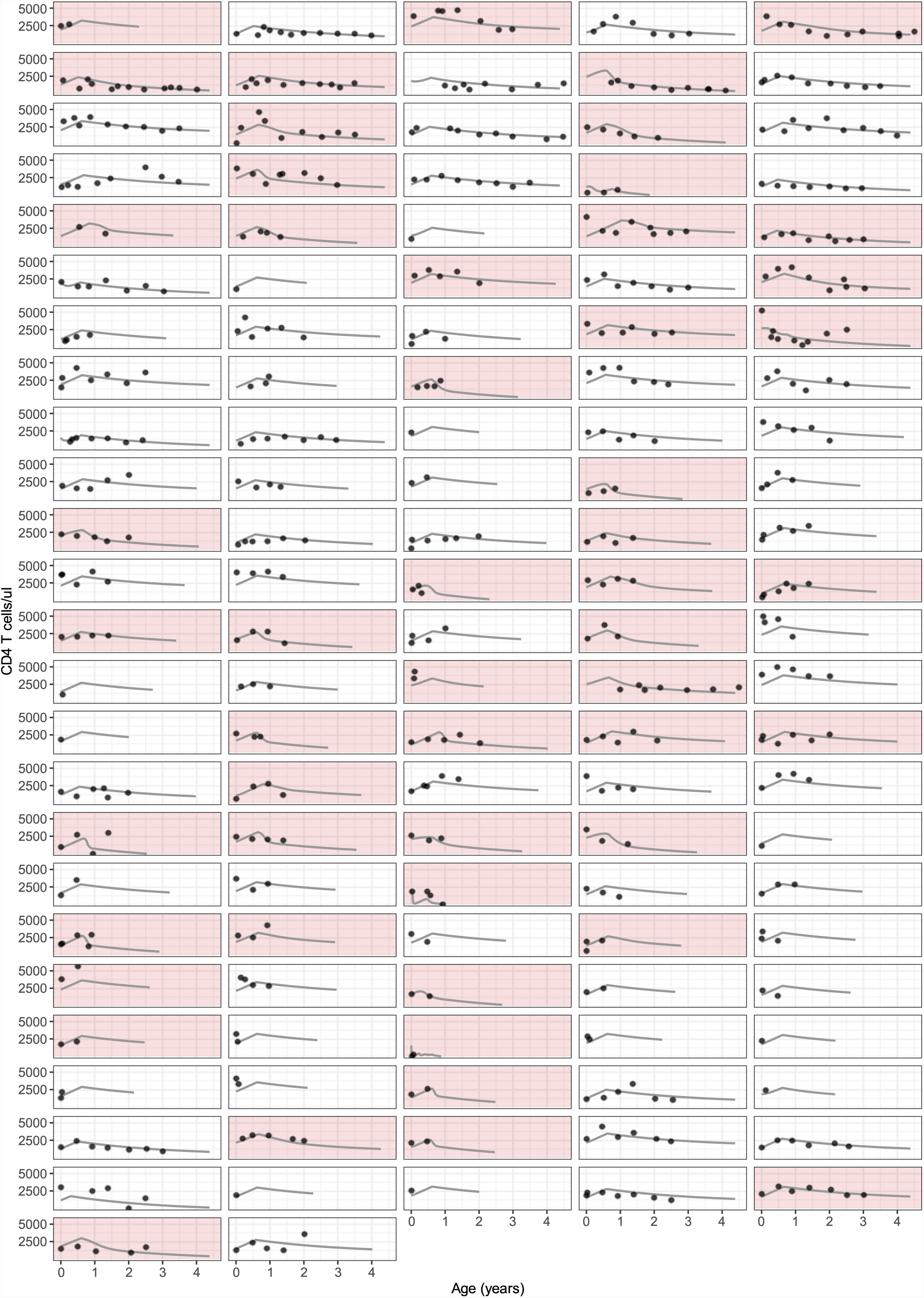
Model fits for CD4 T cell observations. Each panel represents a different infant, ordered as in Fig 2; points represent the data; and solid lines are the model fits.

The fixed effects for all estimated parameters, and the standard error of the associated random effects, are given in Table 1. The population-level average lifespan of productively infected cells (1*/d*) was 19 days (95% percentile across all infants = 5–52 days), and *R*_0_ at ART initiation was 0.35 (0.18–1.25), reflecting an initial decrease in VL across most infants. The rate of CD4 T cell reconstitution, *r*, was positively correlated with the initial number of CD4 T cells, *T*_0_, and the duration of reconstitution, *T*_*R*_ (Fig S7). These associations are unlikely to be driven by poor parameter identifiability, which would instead cause negative correlations through compensatory mechanisms. Notably, we found that infants with higher reconstitution rates, *r*, and VL production to decay ratios, 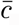, were more likely to experience increases in VL after ART initiation (*p* < 1 ×10^−4^; Fig 4A–B), although the effect was small in the former case (difference in means = 1.2% of the population average). For those infants who did experience increases in VL, larger and earlier increases were associated with higher VL production to decay ratios (*p* < 1 × 10^−4^; Fig 4C–D), but not reconstitution rates (*p >* 0.5). Overall, these results suggest that VL production to decay ratios and CD4 reconstitution rates are the most important parameters determining an infant’s resurgence characteristics.

**Figure 4:**
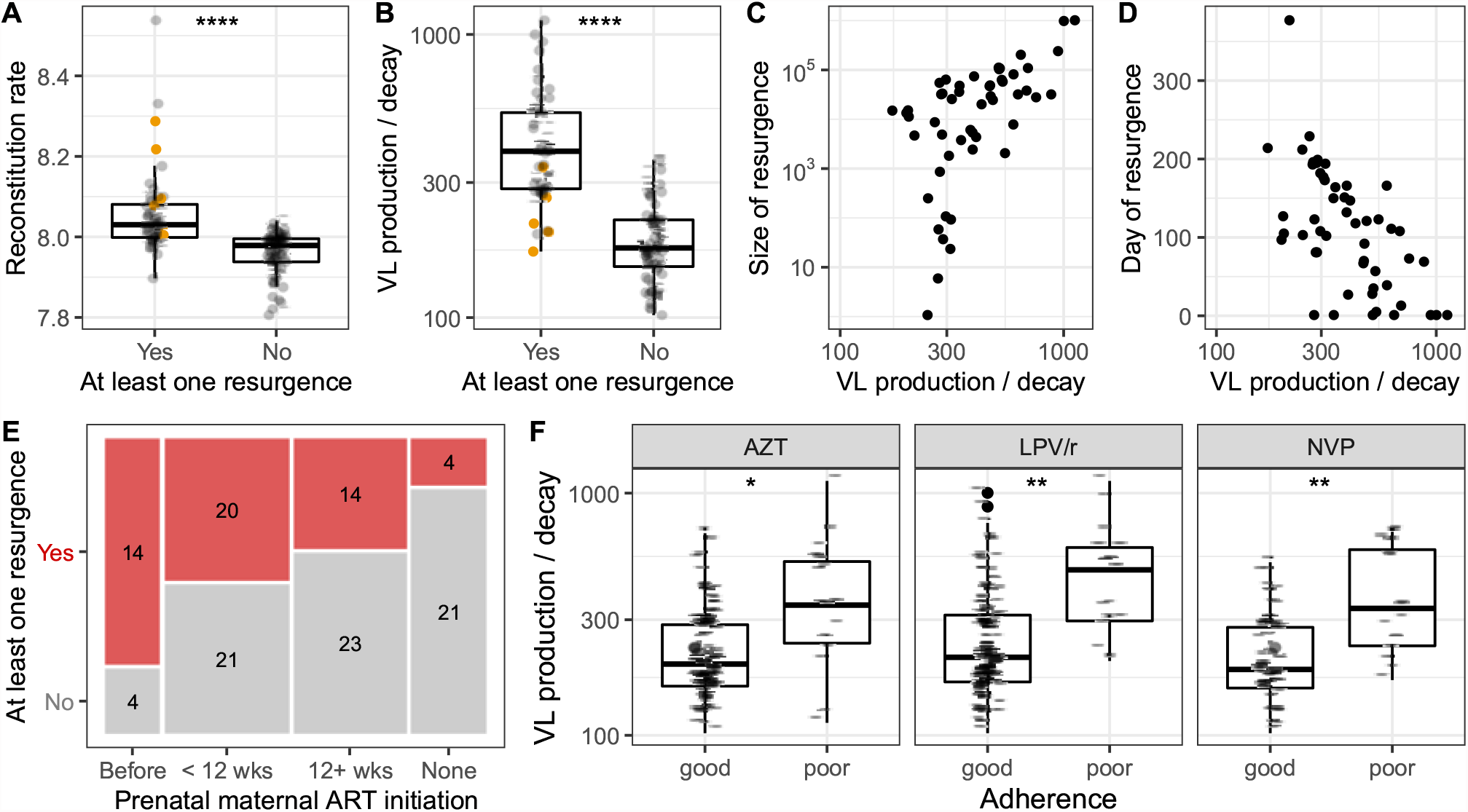
VL resurgence is associated with CD4 reconstitution, VL production and decay, and ART history of infant and mother. (A–B) Relationship between the occurrence of VL resurgence (defined as any increase in VL following initiation of ART) and the CD4 reconstitution rate, *r*, in cells *µ*l^-1^ day^-1^ (A) and ratio of VL production to decay, 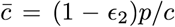, in copies ml^-1^ cell^−1^ (B). Each point represents a different infant and *p* < 0.0001(^∗∗∗∗^) in both cases. Five infants whose resurgence was a viral rebound event are highlighted in orange. (C–D) Relationship between 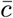 and the size of VL resurgence in RNA copies ml^-1^ (C) and timing of VL resurgence in days (D). Each point represents an infant who experienced resurgence. Correlations are 0.61 and −0.58, respectively, and *p* < 1× 10^−4^ in both cases. (E) Relationship between the occurrence of VL resurgence and the timing of maternal prenatal ART initiation (*p* < 0.01). The size of each box reflects the proportion of infants in the corresponding category and the numbers show the corresponding sample size. (F) Relationship between infant ART adherence is associated and VL production to decay ratios. Adherence was classified as ‘good’ if the majority of adherence estimates were 90% or more, and ‘poor’ otherwise. The VL production to decay ratio is given by 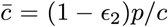, in copies ml^-1^ cell^−1^. Each panel represents a different drug and each point represents a different infant. Significance levels are *p* < 0.05(^∗^); *p* < 0.01(^∗∗^).

In addition to the deterministic model parameters, we found a longer duration of maternal prenatal ART was associated with risk of VL resurgence (*p* < 0.01; Fig 4E). However, this covariate was also associated with higher VL production to decay ratios (*p* < 0.01; Fig S9), suggesting potential colinearity. All other associations between VL resurgence characteristics and clinical covariates, including pre-treatment CD4 percentage and age at ART initiation (Table S1), were not significant at the *p* = 0.05 level. Taken together, these findings suggest maternal prenatal ART history may mediate the risk of VL resurgence, through, or in addition to, the deterministic model parameters discussed above.

### Variation in both adherence and the natural dynamics of CD4 T cells dictate infant trajectories

The most obvious explanation for the wide variety of VL suppression and resurgence trajectories we have identified here is variation in ART adherence, which may be more pronounced in infants than adults. In our models, we assume variation in ART adherence is reflected entirely in the parameters *ϵ*_1_ and/or *ϵ*_2_, which dictate the efficiency of treatment at blocking new infection and virus production by infected cels, respectively. These parameters are not individually identifiable with these data, but instead are subsumed in the compound parameter 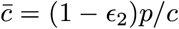 and *β*_0_ = (1 − *ϵ*_1_)*β*. (In support of this, we found that 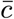 was associated with our measures of LPV/r, NVP and AZT adherence from the study questionnaires (*p*<0.05, Fig 4F), although *β*_0_ was not.) Thus, if adherence is the main driver of variation in VL trajectories, then individual variation in 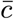 and *β*_0_ should be the most crucial components of our model. However, the association we detected between CD4 reconstitution rates and the probability of resurgence suggest variation in CD4 T cell dynamics may also be important. To explore this issue, we refit the model while removing the random effects for *β*_0_, 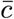 and *r* in turn. We found that fixing any of these parameters resulted in poorer fits, although the model with fixed *r* performed worst overall (Table 3). This suggests that CD4 T cell dynamics are an important factor driving variation in HIV suppression and resurgence characteristics across infants, in addition to ART adherence.

**Table 3:** Model comparisons of adherence and CD4 recovery parameters. AIC values (ΔAIC) are quoted relative to the minimum AIC value across all models. The model with ΔAIC = 0 is the model with lowest AIC and thus has most statistical support. See Table 1 for parameter definitions.

| Model | $\Delta\text{AIC}$ |
| --- | --- |
| Fixed and random effects for $\beta_0$, $\bar{c}$ and $r^*$ | 0.0 |
| Fixed and random effects for $\bar{c}$ and $r$; only fixed effects for $\beta_0$ | 30.0 |
| Fixed and random effects for $\beta_0$ and $r$; only fixed effects for $\bar{c}$ | 31.9 |
| Fixed and random effects for $\beta_0$ and $\bar{c}$; only fixed effects for $r$ | 79.6 |

\*Corresponds to the best-fit model in Table 2.

## Discussion

In this study we modeled the dynamics of HIV suppression and rebound in perinatally-infected infants receiving ART. Our framework extends previous models of rebound in adults [7, 12] by incorporating mechanisms of the natural decline and infection-induced reconstitution of CD4 T cells in young infants [14]. We found that new infection events were initially the major contributor to growth of the productively infected cell population, but that reactivation of the latent reservoir became more important once VL levels were low (Fig S4). We also identified natural processes as the longterm driver of CD4 T cell declines in blood.

Although our estimates of the average CD4 T cell reconstitution rate (*r* = 8 cells *µ*l^-1^ day^-1^) is greater than those from another cohort of HIV-infected infants (*r* = 3.8 cells *µ*l^-1^ day^-1^), it is within the interquartile ranges. Notably, infants from this other cohort initiated ART later, on average, than the infants in our cohort (median = 82 days, IQR = 34–121), and all eventually achieved viral suppression [14]. We also found that higher rates of reconstitution were associated with a greater probability of experiencing a resurgence in VL. This relationship was not confounded by the immunological status of infants at the beginning of the study as we found no association between the reconstitution rate and pre-treatment CD4 percentage or counts, or between the risk of VL resurgence and pre-treatment CD4 percentage or counts. Our finding raises the possibility that rapid recovery of CD4 T cells, despite suggesting an improved clinical state, can also increase the risk of VL resurgence in some individuals by repopulating the target cell pool. Although it could also be that VL resurgence triggers more rapid CD4 reconstitution through increased anti-viral immune activity or density-dependent responses to CD4 depletion [14], the latter seems unlikely in this cohort as we did not detect a negative association between the initial number of CD4 T cells (*T*_0_) and *r*. Nevertheless, further investigation is needed to determine the directionality of this relationship, and whether the extent of CD4 T cell recovery may be used as a biomarker for individuals at increased risk of VL resurgence.

We also found that higher ratios of VL production to decline, 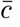, were associated with a greater risk of resurgence, and larger and earlier increases in VL given that resurgence occurred. This is perhaps to be expected given that 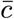 effectively controls the amount of free virus available to infect new cells at any given time. The association between 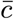 and LPV/r adherence is also expected given that 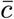 is a function of the protease inhibitor efficacy, *ϵ*_2_. However, the additional associations with AZT and NVP adherence suggest 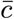 is also capturing some of the variation in reverse transcriptase inhibitor adherence through virus availability and its downstream effects on cell infection rates. Finally, although we cannot isolate the contribution of drug resistance to the protease inhibitor efficacy, we expect that resistance to LPV/r is rare in this cohort and is thus unlikely to be a major driver of individual variation in 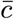, or of VL resurgence patterns.

Our estimate of another key parameter, the rate of latent cell reactivation (*a* = 10^−3^ cells *µ*l^-1^ day^-1^), is at the upper limit of similar estimates from adults obtained during ART interruption (2 × 10^−6^ – 1 × 10^−3^ cells *µ*l^-1^ day^-1^ [7]). Biologically, a higher burden of reactivation (*a*) may reflect a larger latent reservoir in these infants and/or an increased per-cell rate of latent cell reactivation. Dynamically, larger reactivation estimates may compensate for significant fluctuations in treatment adherence that are not included in the model. We did not find any associations between *a* and the occurrence or size of VL resurgence. This is not surprising as *a* effectively represents the total contribution of latent cell reactivation averaged over the entire study, and its contribution to changes in VL relative to those of de novo infection events is small (Eqn. 2).

One unexpected result is that our simple framework can capture large variations in infant VL trajectories, including monotonic decreases to sustained suppression, resurgences in VL, and suppression with subsequent rebound. Although the canonical explanation for erratic VL patterns is imperfect ART adherence, we found that incorporating variation in CD4 reconstitution rates was also required to capture the complexity in our infant data. In addition, we found that longer durations of maternal prenatal ART were associated with VL resurgence, which is consistent with findings from this cohort that exposure to maternal prenatal ART is associated with a larger viral reservoir [29], and could not be explained by worse adherence in this group (*p >* 0.2). However, we could not disentangle the effects of this variable from that of VL production to decay ratios, 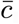. Netherless, our findings demonstrate that a deterministic framework with reactivation and constant ART efficacy can recapitulate intricate infection dynamics, and that, with the levels of adherence achieved in this study, resurgence may in fact be inevitable for infants with certain virological and CD4 T cell parameter combinations.

There are a number of caveats to our modeling approach. First, our model does not differentiate between shortand long-lived productively infected cells, the loss of which underpins the multiphasic decline of VL in adults and infants on ART [4, 30, 31], or cells that have undergone abortive infection [17]. Instead, our estimates of the mean lifespan of an infected cell is effectively a weighted average of the mean lifespans of these subpopulations. Although our averaged estimate (19 days) is longer than corresponding values in adults (0.5–6 days) [7], it agrees with the median short and long-lived cell lifespans we estimated previously in a subset of these infants who achieved suppression (17 days) [4]. Second, we do not explicitly model the dynamics of latently infected cells as they are not directly observed. However, the parameter *a* in our model is effectively a ‘force of reactivation’, which combines the effects of reservoir size and the per-cell rate of reactivation. Third, we do not explicitly model non-productively infected cells. Although observed VL is likely a combination of infectious and non-infectious virus, our results will hold if the ratio of these remains approximately constant over time for each infant. Substantial and sustained changes in LPV/r efficacy (*ϵ*_2_), for example, through prolonged shifts in adherence levels and/or increasing drug resistance, may cause non-negligible changes in this ratio. However, fluctuations in LPV/r adherence, although frequent, were usually transient (Fig S12), and we expect LPV/r resistance in this cohort to be rare. Nevertheless, a potentially important extension of our model would be the inclusion of non-productively infected cells and/or allowing changes in LPV/r efficacy over time. Lastly, we fit the peripheral CD4 T cell data to the number of target cells predicted by the model (*T* (*t*)), rather than the predicted sum of target cells, productively infected cells and latently infected cells. This approach is reasonable as the frequency of infection in CD4 T cells is usually small [32], and the majority of infected cells most likely reside in lymphoid tissues where infection-induced CD4 depletion is greatest [33]. We also assume all CD4 T cells are equally susceptible to infection, although in reality activated cells may be more susceptible than resting cells [34, 35]. However, this heterogeneity is implicitly incorporated within the transmission parameter, *β*, if the proportion of CD4 T cells that are susceptible remains approximately constant over time.

Finally, we acknowledge that the ART regimens used in the LEOPARD trial may not be optimal. Although considered most effective at the time of study design and implementation, more potent treatments – for example, integrase inhibitors and/or broadly neutralizing antibodies – have since been approved for young infants. It will be important to determine whether infants starting these newer treatments are also at risk of the inevitable resurgence we have identified here.

In conclusion, we have extended the classic framework for HIV suppression and rebound to include more realistic dynamics of CD4 T cell decline and reconstitution in young infants on ART. We estimated rates of reactivation and reconstitution, and identified distinct phases in which dynamics were either dominated by new infection of CD4 T cells, or by reactivation of the latent reservoir. Moreover, we demonstrated the importance of incorporating variation in CD4 reconstitution rates to capture the diversity of infant VL trajectories. Overall, our results suggest that VL resurgence in perinatally-infected infants may be inevitable in certain parameter regimes, and highlight the utility of mathematical modeling in understanding the dynamics of infant HIV infection.

## Data Availability

All data produced in the present study are available upon reasonable request to the authors

## Acknowledgements

This work is part of the EPIICAL project (http://www.epiical.org/), supported by the PENTA-ID foundation (http://penta-id.org/), funded through an independent grant by ViiV Healthcare UK. Data were collected during the Latency and Early Neonatal Provision of Antiretroviral Drugs Clinical Trial (LEOPARD) study. The LEOPARD study was supported in part by the Eunice Kennedy Shriver National Institute of Child Health and Human Development/National Institute of Allergy and Infectious Disease, National Institutes of Health (U01HD080441), USAID/PEPfAR, the South African National HIV Programme, and South African Research Chairs Initiative of the Department of Science and Technology and National Research Foundation of South Africa.

## Latency and Early neOnatal Provision of Anti-Retroviral Drugs (LEOPARD) Study Team

Louise Kuhn, Elaine Abrams, Wei-Yann Tsai, Stephanie Shiau (Columbia University Medical Center, New York); Caroline Tiemessen, Maria Paximadis, Sharon Shalekoff, Diana Schramm, Gayle Sherman (National Institute of Communicable Diseases (NICD), Johannesburg, South Africa); Renate Strehlau, Megan Burke, Martie Conradie, Ashraf Coovadia, Ndileka Mbete, Faeezah Patel, Karl Technau (Empilweni Services and Research Unit (ESRU), Rahima Moosa Mother and Child Hospital, Johannesburg, South Africa); Grace Aldrovandi (University of California, Los Angeles); Rohan Hazra (Eunice Kennedy Shriver National Institute of Child Health and Human Development); Devasena Gnanashanmugam (National Institutes of Allergy and Infectious Diseases).

## The EPIICAL Consortium study team

Nigel Klein, Diana Gibb, Sarah Watters, Man Chan, Laura McCoy, Abdel Babiker (University College London, UK); Anne-Genevieve Marcelin, Vincent Calvez (Université Pierre et Marie Curie, France); Maria Angeles Munoz (Servicio Madrileño de Salud-Hospital General Universitario Gregorio Marañon, Spain); Britta Wahren (Karolinska Institutet, Sweden); Caroline Foster (Imperial College Healthcare NHS Trust, London, UK); Mark Cotton (Stellenbosch University-Faculty of Medicine and Health Sciences, South Africa); Merlin Robb, Jintanat Ananworanich (The Henry M. Jackson Foundation for the Advancement of Military Medicine, Maryland); Polly Claiden (HIV i-Base, UK); Deenan Pillay (University of KwaZulu-Natal Africa Center, South Africa); Deborah Persaud (Johns Hopkins University); Rob J de Boer, Juliane Schröter, Anet J N Anelone (University of Utrecht, Netherlands); Thanyawee Puthanakit (Thai Red Cross AIDS-Research Centre, Thailand); Adriana Ceci, Viviana Giannuzzi (Consorzio per Valutazioni Biologiche e Farmacologiche, Italy); Kathrine Luzuriaga (University of Massachusetts Medical School, Worcester, Massachusetts); Nicolas Chomont (Centre de Recherche du Centre Hospitalier de l’Université de Montreal-University of Montreal, Canada); Mark Cameron (Case Western Reserve University, Cleveland, Ohio); Caterina Cancrini (Università degli Studi di Roma Tor Vergata, Italy); Andrew J Yates, Louise Kuhn, Sinead E Morris (Columbia University Medical Center, New York); Avy Violari, Kennedy Otwombe (University of the Witwatersrand, Johannesburg [PHRU], South Africa); Ilaria Pepponi, Francesca Rocchi (Children’s Hospital “Bambino Gesu”, Rome, Italy); Stefano Rinaldi (University of Miami, Miller School of Medicine, Florida); Alfredo Tagarro (Hospital 12 de Octubre, Universidad Complutense, Madrid, Spain); Maria Grazia Lain, Paula Vaz (Fundação Ariel Glaser contra o SIDA Pediátrico, Mozambique); Elisa Lopez, Tacita Nhampossa (Fundação Manhiça, Mozambique).

We thank Juliane Schröter and Rob de Boer for critical reading and helpful suggestions.

## Supplementary Information

### Additional information on infant adherence

At each study visit, the infant’s caregiver provided additional information for a questionnaire taken by the attending physician. Caregivers were asked if any doses had been missed since the previous visit and, if so, how many. They were also asked about any challenges administering the medication, including drug tolerance issues (e.g. if the infant spit up the medicine and repeat doses were required).

This information was a valuable supplement to estimates of adherence calculated from the amount of medication returned at the visit, given that these were often missing (due to leftover medication spilling or being left at home). If a particular adherence estimate was missing, we checked the corresponding questionnaire; if the physician noted serious adherence concerns for that drug, such as a series of missed doses, adherence was labeled as ‘poor’. See Fig S10 for the resulting time series.

### Model

Starting with the original set of equations

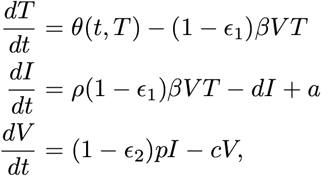

we assume that viral dynamics occur on a faster timescale than those of CD4 T cells, i.e. *dV/dt* = 0. This gives *I* = *cV /*(1 *− ϵ*_2_)*p*, and we can rewrite the above equations as

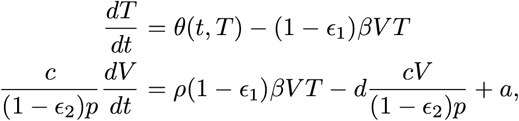

i.e.

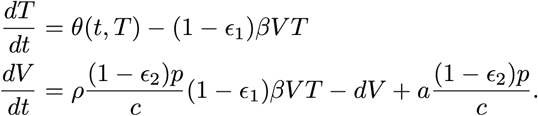

Setting *β*_0_ = (1 *− ϵ*_1_)*β* and 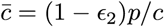 then gives the reduced system,

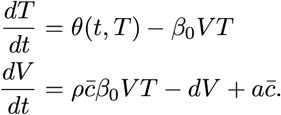

### Structural Identifiability

We explore the structural identifiability of the equations using the approach of Castro and de Boer (2020) [1]. First, we define scaling factors for all parameters we want to estimate, i.e. 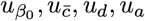, and 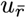. The scaling factors for all fixed parameters are equal to one. Similarly, since both *V* and *T* are observed, we do not need to define any variable scaling factors.

Next, we equate all functionally independent terms in our equations that contain these parameters to their scaled counterparts:

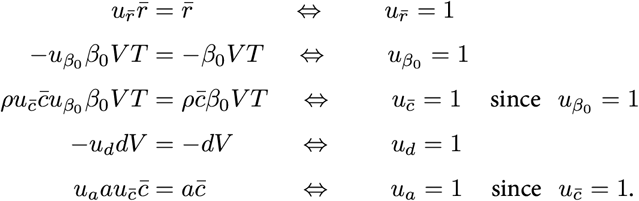

Since all scaling factors have solution equal to 1, all estimated parameters are identifiable. Note that we assume *r* is identifiable if 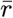 is identifiable and *T*_*R*_ is fixed.

### Nonlinear Mixed Effects Modeling in Monolix

We fit the following system of equations in Monolix,

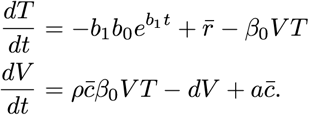

All VL observations below the detection threshold of 20 copies ml^−1^ are treated as censored values. In line with previous pharmacokinetic and viral dynamics modeling, we assumed both *V* (*t*) and *T* (*t*) were lognormally distributed with combined error models, i.e. for variable *X*_*i*_(*t*), the residual error is expressed as the sum of a constant term and a term proportional to *X*_*i*_(*t*) (‘combined1’ in Monolix) [2–4].

We fix *b*_0_, *b*_1_ and *ρ* across infants to the values given in Table 1. Initially we fix *T*_*R*_ = 190 days across all infants, but subsequently explore fits when this value is varied, and when it is freely estimated. All other parameters were estimated and assumed to have both fixed and random effects. Guided by exploratory fits, each estimated parameter was assumed to follow a lognormal distribution, with the exception of *a* which followed a logit-normal distribution between 0 and 0.1, and *r* which followed a normal distribution, and *T*_*R*_ which also followed a normal distribution (when estimated). Initial estimates for all population parameters are given in Table S3. Initial estimates for the residual error models were kept at their default values. Following exploratory fits, we allowed for a correlation between *β* _0_ and *d*, but assumed all other parameters were independent.

### Supplementary Figures and Tables

**Table S1:** Summary of the clinical covariates included in our analyses.

| Covariate | Group | N | Variable treatment |
| --- | --- | --- | --- |
| Sex | Male | 59 | Categorical |
|  | Female | 63 |  |
| Preterm | Yes | 88 | Categorical |
|  | No | 103 |  |
| Delivery mode | Normal vaginal delivery | 87 | Categorical |
|  | Caesarean section | 35 |  |
| Birth weight (g) | <2500 | 29 | Continuous |
|  | 2500+ | 93 |  |
| Age at ART initiation (days) | <2 | 49 | Categorical |
|  | 2–14 | 56 |  |
|  | 14+ | 17 |  |
| Pre-treatment CD4 percentage | <35 | 33 | Continuous |
|  | 35+ | 61 |  |
|  | Not recorded | 28 |  |
| Mother's viral load (copies ml<sup>-1</sup>) | <1000 | 32 | Continuous |
|  | 1000+ | 90 |  |
| Mother's CD4 count (cells $\mu\text{l}^{-1}$) | <350 | 61 | Continuous |
|  | 350+ | 61 |  |
| Mother's CD4 percent | <25 | 77 | Continuous |
|  | 25+ | 45 |  |
| Maternal prenatal ART history | None | 25 | Categorical |
|  | Initiated 12+ weeks into pregnancy | 37 |  |
|  | Initiated <12 weeks into pregnancy | 41 |  |
|  | Initiated before pregnancy | 18 |  |
|  | Unknown | 1 |  |
| Breastfeeding | Some | 96 | Categorical |
|  | None | 26 |  |

**Table S2:** Comparing models with *T*_*R*_ fixed across all infants. The AIC difference for model *i* was calculated as AIC_*i*_ AIC_*min*_, where AIC_*min*_ is the minimum AIC value across all models. The model with zero difference is the model with lowest AIC and thus is the most strongly favored.

| Fixed value of $T_R$ (days) | AIC difference |
| --- | --- |
| 170 | 38.6 |
| 190 | 34.8 |
| 210 | 25.9 |
| 215 | 23.6 |
| 220 | 10.5 |
| 223 | 8.3 |
| **225** | **0.0** |
| 227 | 24.4 |
| 230 | 13.2 |
| 240 | 30.3 |
| 250 | 22.8 |
| 270 | 30.8 |
| 300 | 43.2 |

**Table S3:** Initial estimates for the population parameters in Monolix.

| Parameter | Distribution | Fixed effect | Standard Deviation<br>of Random Effects |
| --- | --- | --- | --- |
| $\beta_0$ | Lognormal | $1 \times 10^{-6}$ | 0.1 |
| $\bar{c}$ | Lognormal | 100 | 1 |
| $d$ | Lognormal | 0.3 | 0.1 |
| $a$ | Logitnormal (0–0.1) | 0.01 | 0.1 |
| $r$ | Normal | 9 | 0.25 |
| $T_R^*$ | Normal | 225 | 2 |
| $T_A^*$ | Normal | 365 | 2 |
| $T_0$ | Lognormal | 3500 | 1 |
| $V_0$ | Lognormal | 10,000 | 1 |

\*when estimated

**Figure S1:**
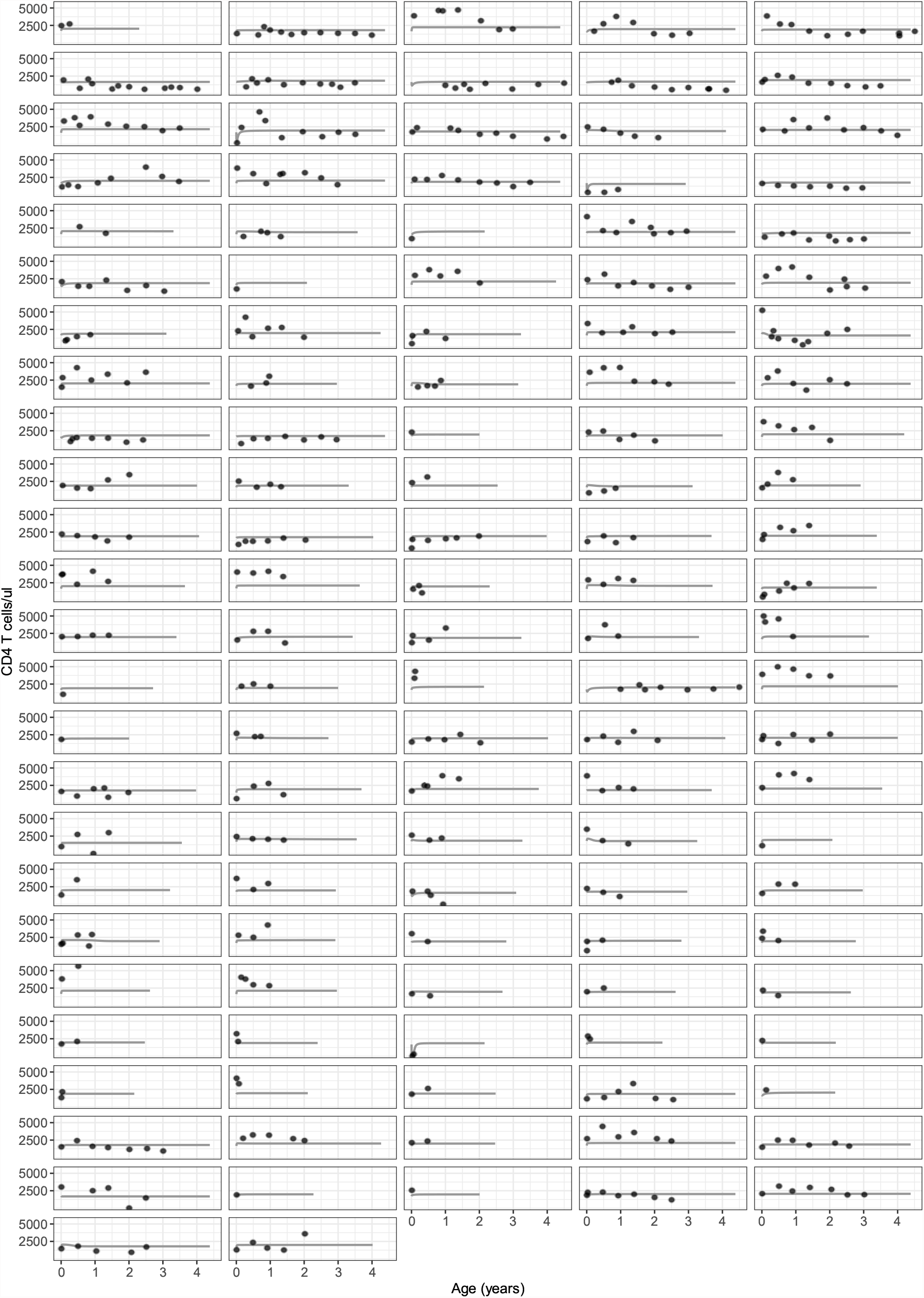
The standard model for adult CD4 T cell dynamics does not capture infant data. Each panel represents a different infant, points represent the data, and solid lines are the model fits. Here *θ*(*t, T*) = *λ − d*_*T*_ *T*, with *λ* and *d*_*T*_ assumed to have lognormal distributions. Initial estimates for the population mean were 1000 cells *µ*l^−1^ day^−1^ and 0.25 day^−1^, respectively, and for the standard deviation were 1 and 0.1, respectively. The nonlinear mixed effects fitting procedure was as described in the main text.

**Figure S2:**
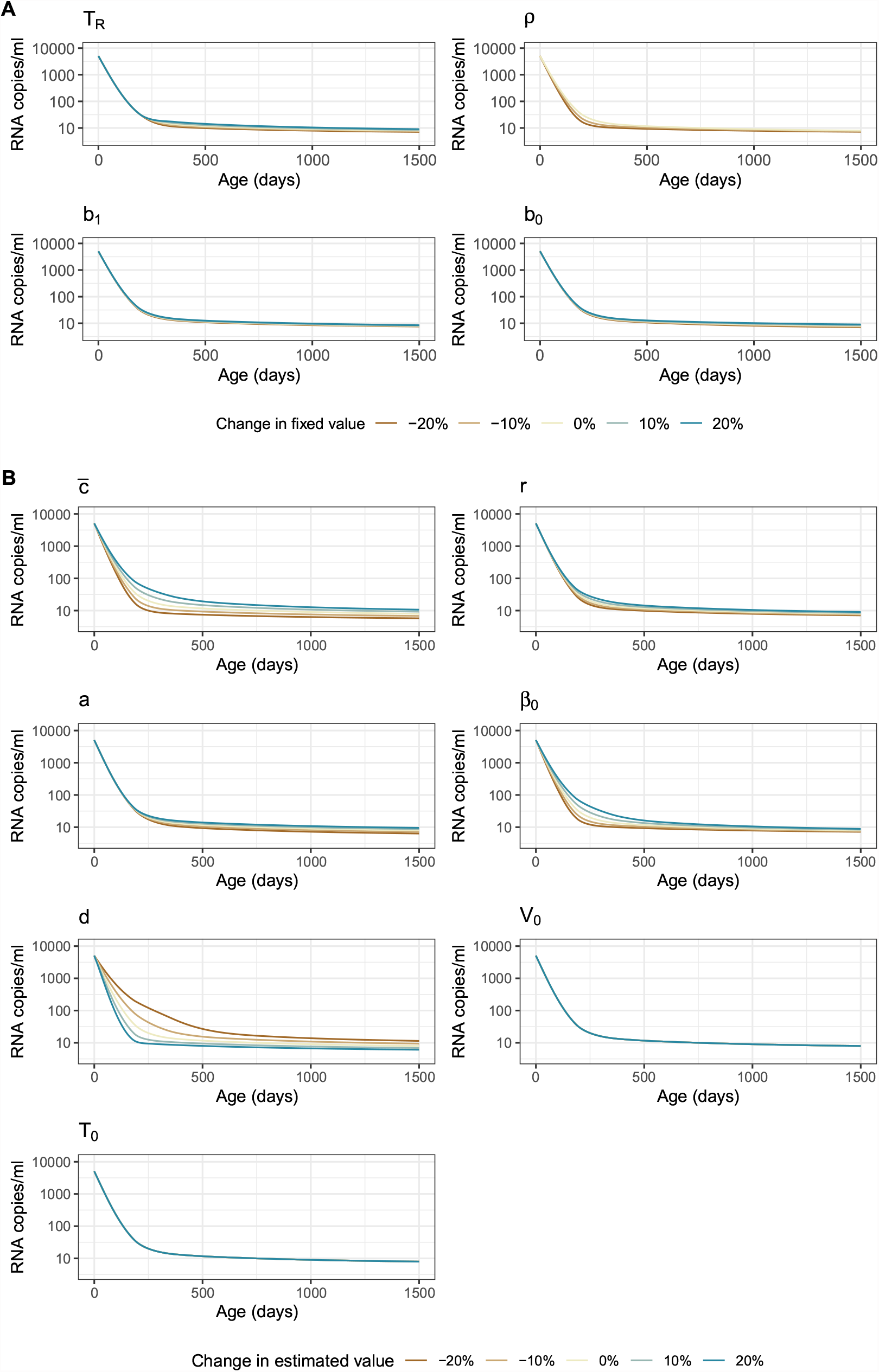
Sensitivity of VL predictions to model parameters. Each fixed (A) or estimated (B) parameter was varied within 20% of its original value while keeping all other parameters at their original values. Original values for the estimated parameters were the population-level means from the best-fit model. Note that predictions for *ρ* do not include 10% and 20% increases as these are biologically unrealistic (i.e. a fraction *>* 1).

**Figure S3:**
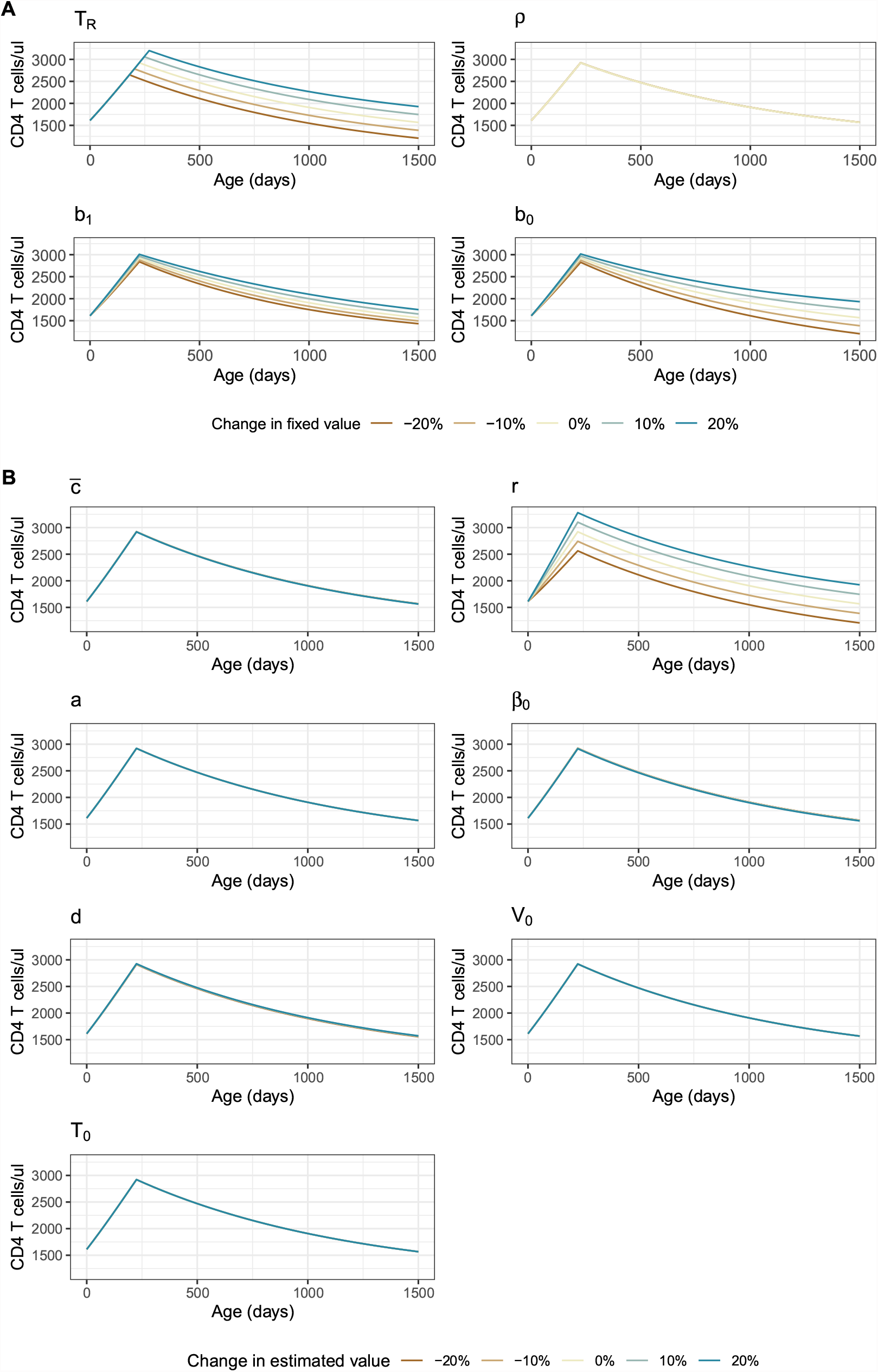
Sensitivity of CD4 T cell predictions to model parameters. Each fixed (A) or estimated (B) parameter was varied within 20% of its original value while keeping all other parameters at their original values. Original values for the estimated parameters were the population-level means from the best-fit model. Note that predictions for *ρ* do not include 10% and 20% increases as these are biologically unrealistic (i.e. a fraction *>* 1).

**Figure S4:**
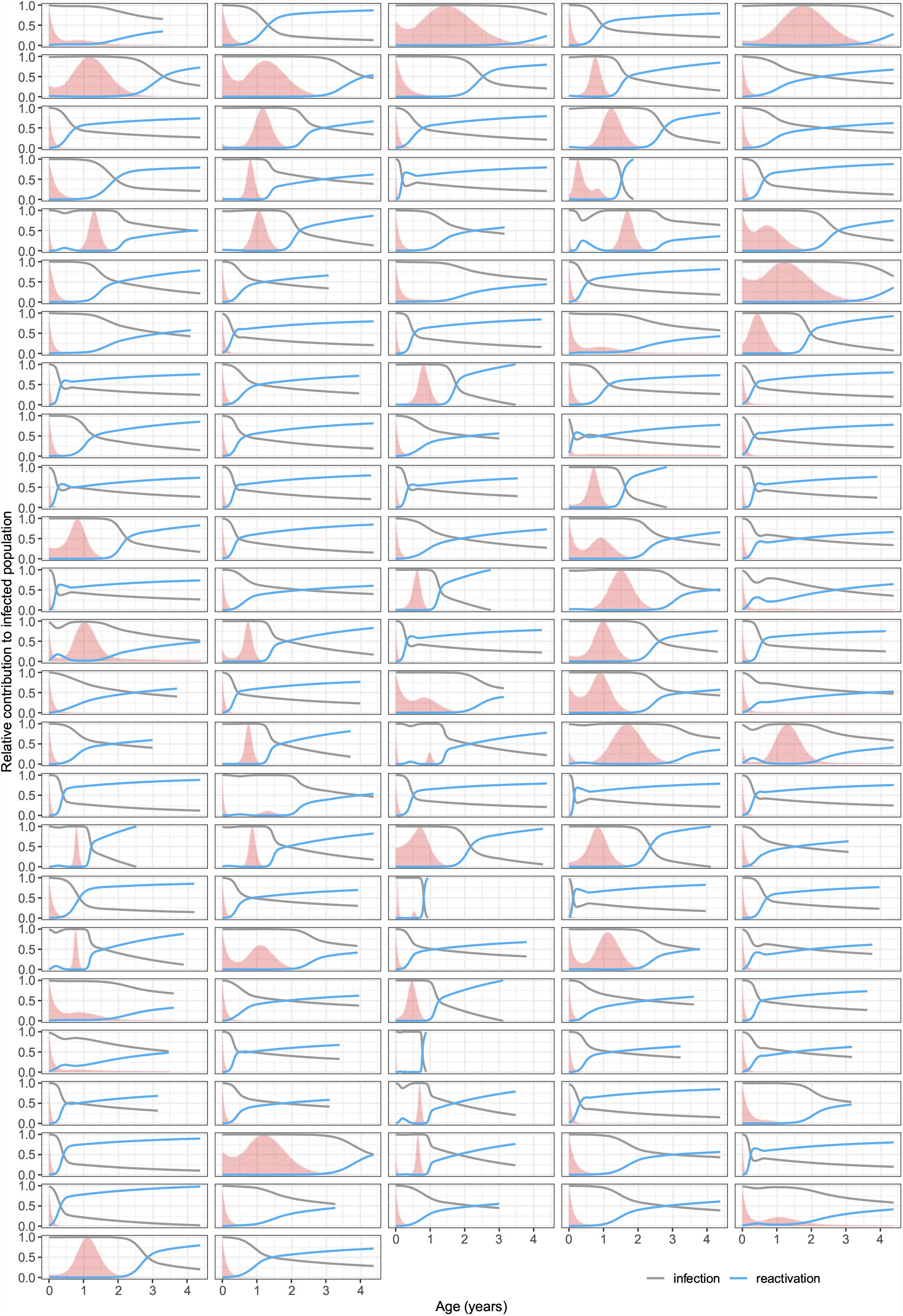
Relative contribution of factors driving an increase in productively infected cells. Each panel represents an infant, and red shaded regions show their VL scaled by its maximum value.

**Figure S5:**
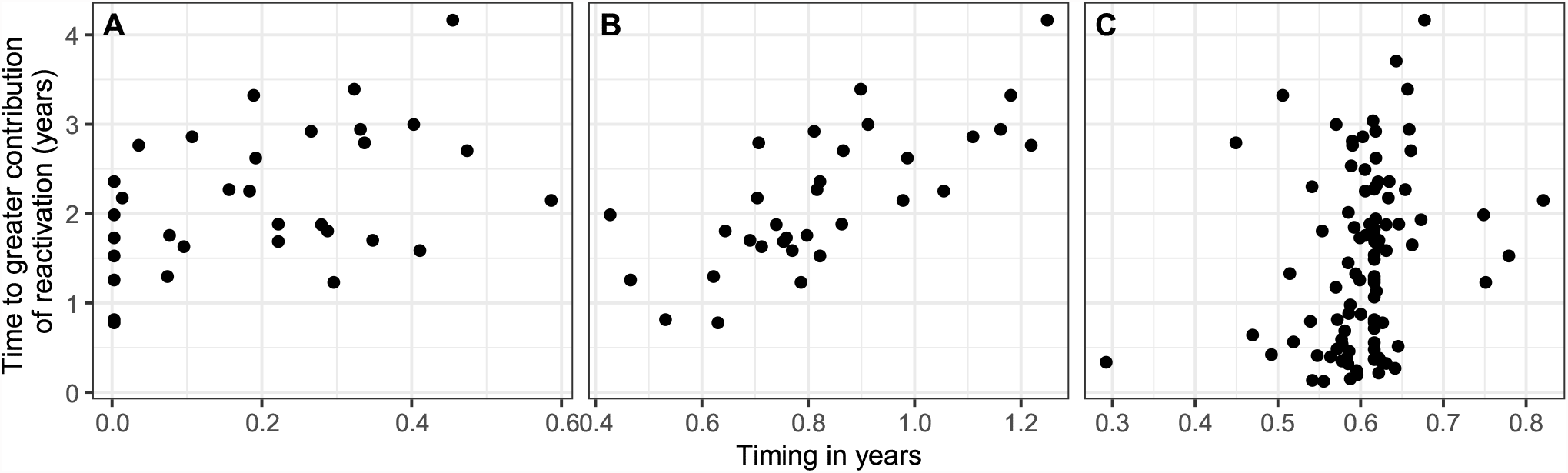
VL and CD4 T cell dynamics influence the time at which reactivation contributes most to productively infected cell growth. Each point represents a different infant with respect to the time at which reactivation became the major contributor to productively infected cell growth and the time at which: (A) their VL started increasing (if applicable); (B) their VL finished increasing (if applicable); and (C) their CD4 T cell recovery plateaued (*T*_*R*_). Correlations are 0.45, 0.71 and 0.29, respectively, and *p* < 0.05 in all cases. Note that the correlation in (C) is reduced to a trend (*p* = 0.05) when the infant with the lowest *T*_*R*_ is removed.

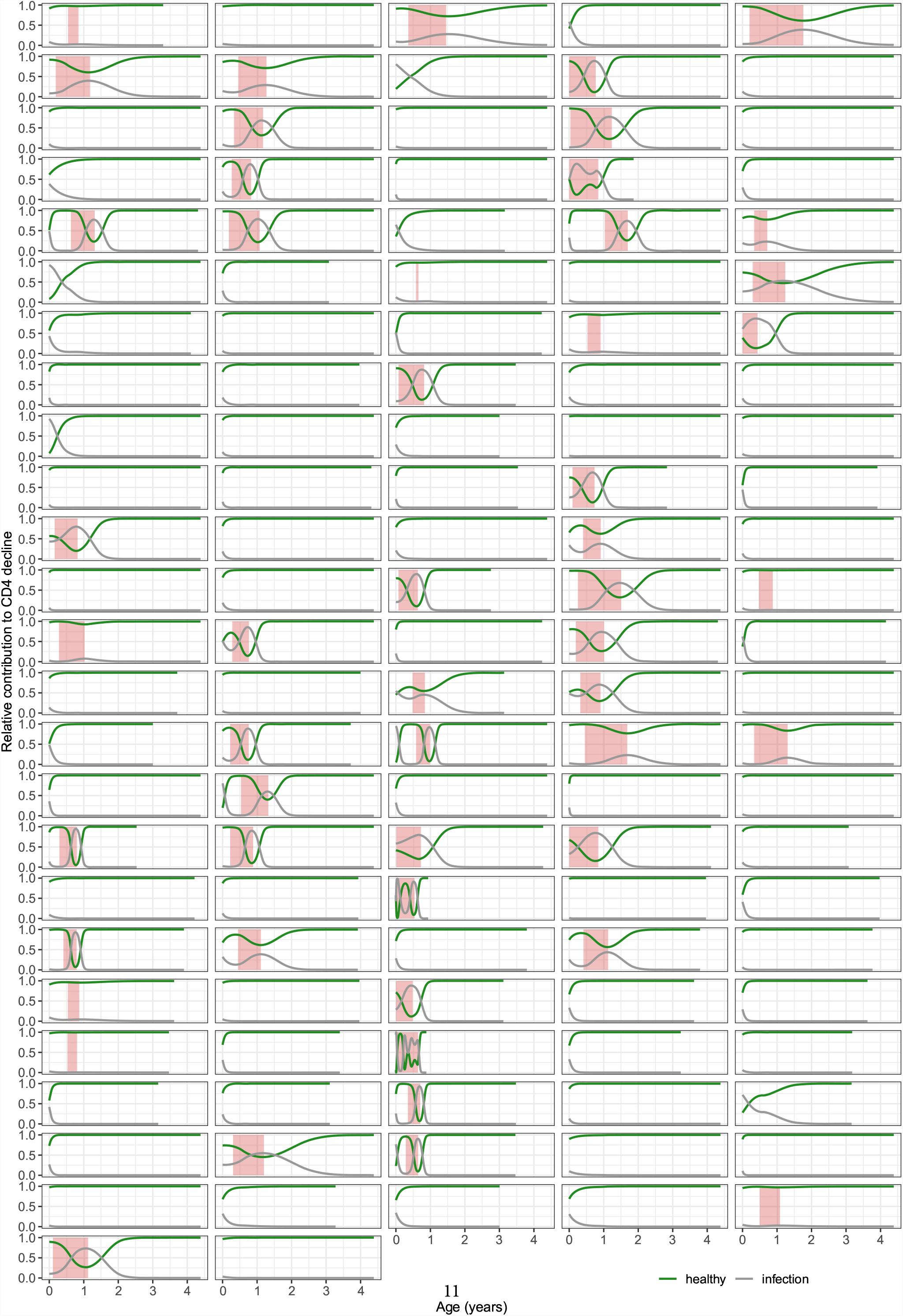

**Figure S7:**
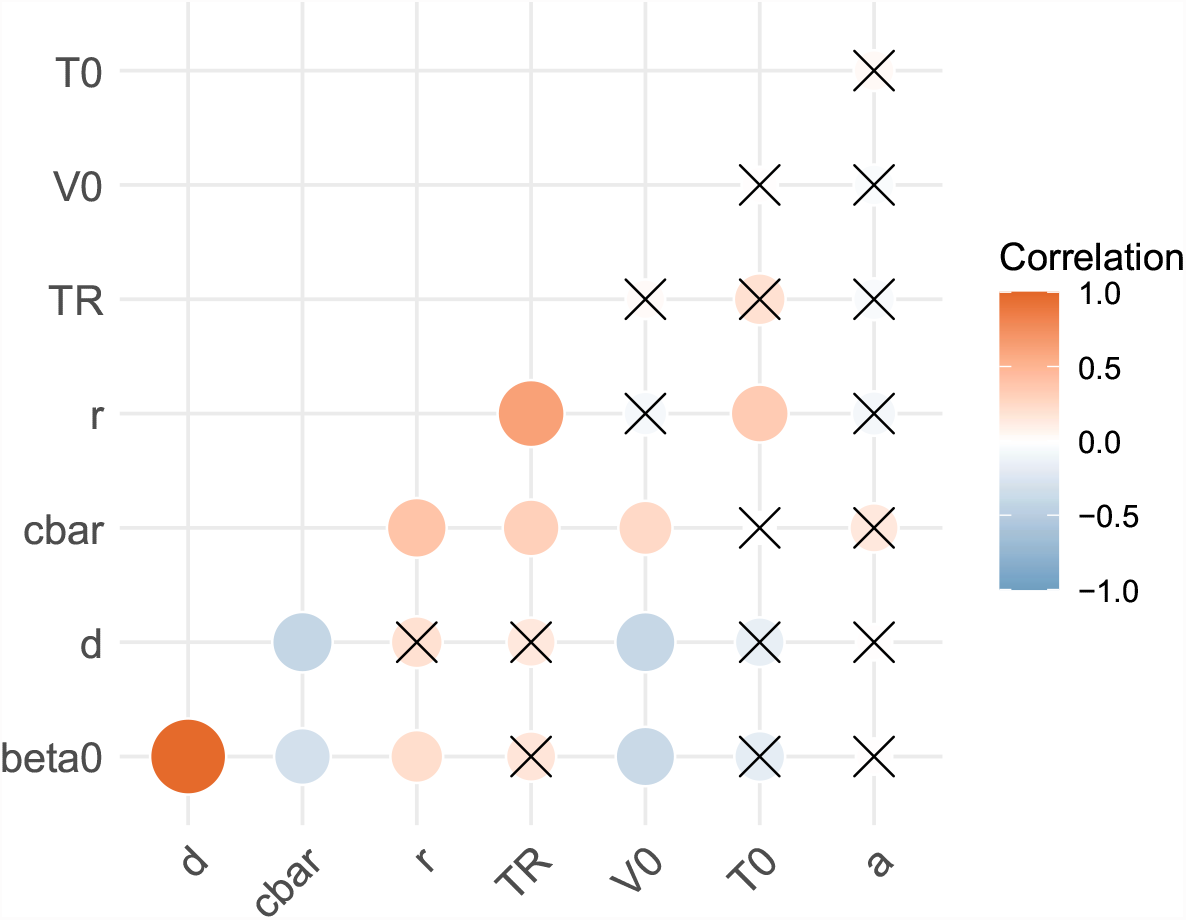
Correlations between estimated parameters. The colour scale shows the strength of the correlation; those with *p*-values greater than a significance threshold of 0.05 are crossed out. *p*-values were adjusted using the Benjamini-Hochberg correction. The strong correlation between *β*_0_ and *d* was included in the nonlinear mixed effects model framework.

**Figure S8:**
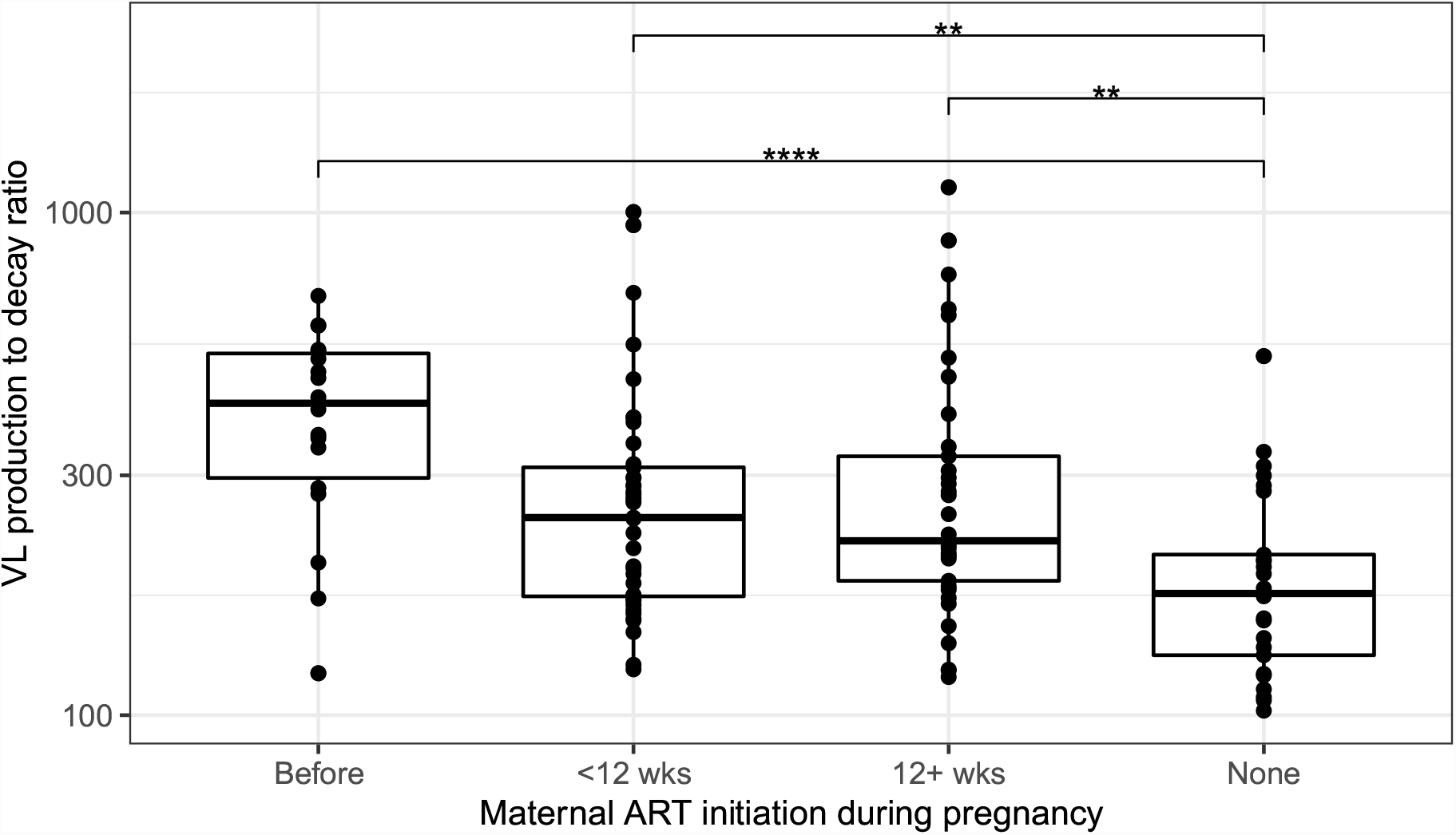
Longer duration of maternal ART is associated with greater VL production to decay ratios. The ratio is given by 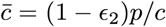, in copies ml^−1^ cell^−1^. Each point represents a different infant. Significance levels are *p* < 0.01(^∗∗^); *p* < 0.001(^∗∗∗^).

**Figure S9:**
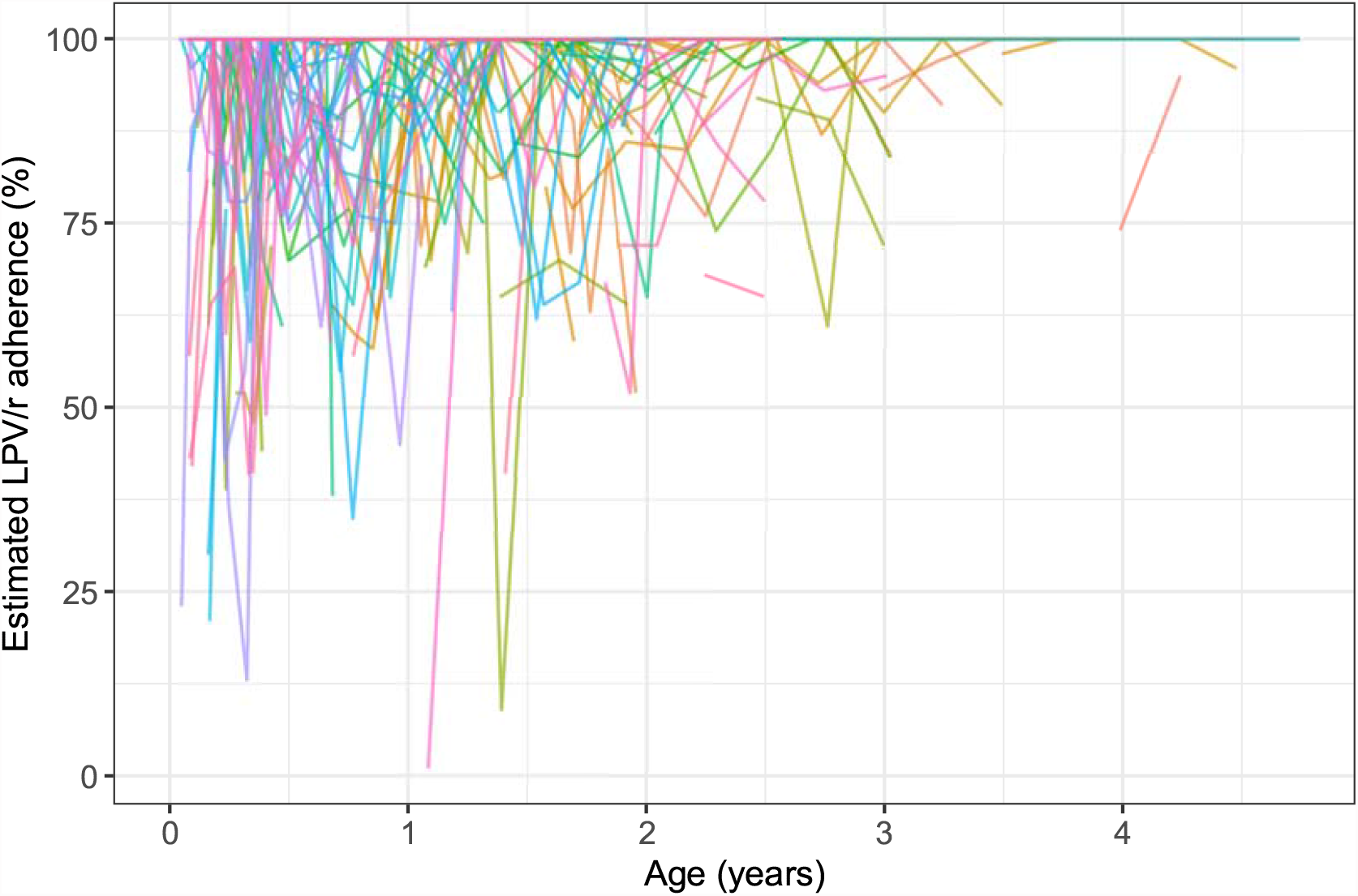
LPV/r adherence estimated from returned medication. Each line represents a different infant.

**Figure S10:**
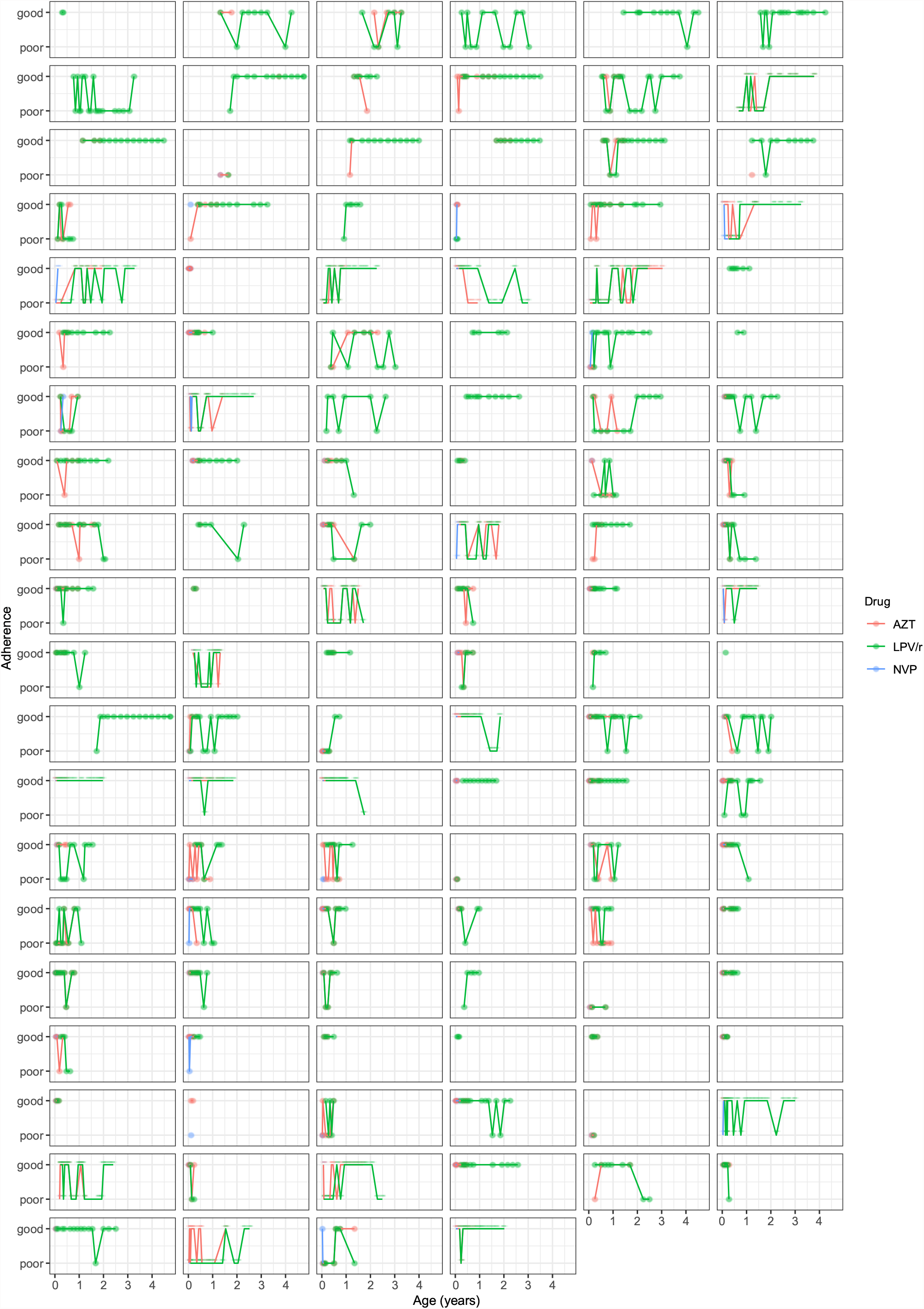
Reported adherence trajectories. Adherence estimates greater than 90% were labeled ‘good’; and all others ‘poor’. Each panel is a different infant.

## References

1. UNAIDS data; 2020. Available from: https://www.unaids.org/en/resources/documents/2020/unaids-data.

2. Luzuriaga K, Gay H, Ziemniak C, et al. Viremic Relapse after HIV-1 Remission in a Perinatally Infected Child. New England Journal of Medicine. 2015;372(8):786–788. doi:10.1056/NEJMc1413931.

3. Violari A, Cotton M, Kuhn L, et al. A child with perinatal HIV infection and long-term sustained virological control following antiretroviral treatment cessation. Nat Comm. 2019;10(412):1–11. doi:10.1038/s41467-019-08311-0.

4. Morris SE, Dziobek-Garrett L, Strehlau R, et al. Quantifying the dynamics of HIV decline in perinatally-infected neonates on antiretroviral therapy. JAIDS. 2020;85(2):209–218. doi:10.1097/QAI.0000000000002425.

5. Kim H, Perelson AS. Viral and Latent Reservoir Persistence in HIV-1–Infected Patients on Therapy. PLOS Computational Biology. 2006;2(10):1–16. doi:10.1371/journal.pcbi.0020135.

6. Archin NM, Vaidya NK, Kuruc JD, et al. Immediate antiviral therapy appears to restrict resting CD4+ cell HIV-1 infection without accelerating the decay of latent infection. Proceedings of the National Academy of Sciences. 2012;109(24):9523–9528. doi:10.1073/pnas.1120248109.

7. Luo R, Piovoso MJ, Martinez-Picado J, et al. HIV Model Parameter Estimates from Interruption Trial Data including Drug Efficacy and Reservoir Dynamics. PLoS One. 2012;7(7):1–12. doi:10.1371/journal.pone.0040198.

8. Hill AL, Rosenbloom DIS, Fu F, et al. Predicting the outcomes of treatment to eradicate the latent reservoir for HIV-1. Proceedings of the National Academy of Sciences. 2014;111(37):13475–13480. doi:10.1073/pnas.1406663111.

9. Petravic J, Rasmussen TA, Lewin SR, et al. Relationship between Measures of HIV Reactivation and Decline of the Latent Reservoir under Latency-Reversing Agents. Journal of Virology. 2017;91(9). doi:10.1128/JVI.02092-16.

10. Reeves DB, Duke ER, Hughes SM, et al. Anti-proliferative therapy for HIV cure: a compound interest approach. Scientific Reports. 2017;7(4011). doi:10.1038/s41598-017-04160-3.

11. Hill AL, Rosenbloom DIS, Nowak MA, et al. Insight into treatment of HIV infection from viral dynamics models. Immunological Reviews. 2018;285(1):9–25. doi:https://doi.org/10.1111/imr.12698.

12. Prague M, Gerold JM, Balelli I, et al. Viral rebound kinetics following single and combination immunotherapy for HIV/SIV. bioRxiv. 2019;doi:10.1101/700401.

13. Huenecke S, Behl M, Fadler C, et al. Age-matched lymphocyte subpopulation reference values in childhood and adolescence: application of exponential regression analysis. European journal of haematology. 2008;80(6):532–539.

14. Schröter J, Anelone AJN, de Boer RJ. Quantification of CD4 recovery in early-treated infants living with HIV. In revision. 2021;.

15. Kuhn L, Strehlau R, Shiau S, et al. Early antiretroviral treatment of infants to attain HIV remission. EClinicalMedicine. 2020;18. doi:10.1016/j.eclinm.2019.100241.

16. Morris SE, Dziobek-Garrett L, Yates AJ. ushr: Understanding suppression of HIV in R. BMC Bioinformatics. 2020;21(52). doi:10.1186/s12859-020-3389-x.

17. Ke R, er Cong M, Li D, et al. On the Death Rate of Abortively Infected Cells: Estimation from Simian-Human Immunodeficiency Virus Infection. Journal of Virology. 2017;91(18):e00352–17. doi:10.1128/JVI.00352-17.

18. Ramratnam B, Bonhoeffer S, Binley J, et al. Rapid production and clearance of HIV-1 and hepatitis C virus assessed by large volume plasma apheresis. Lancet. 1999;354(9192):1782–1785. doi:10.1016/S0140-6736(99)02035-8.

19. Payne H, Lawrie D, Nieuwoudt M, et al. Comparison of Lymphocyte Subset Populations in Children From South Africa, US and Europe. Frontiers in Pediatrics. 2020;8:406. doi:10.3389/fped.2020.00406.

20. Mould D, Upton R. Basic Concepts in Population Modeling, Simulation, and Model-Based Drug Development – Part 2: Introduction to Pharmacokinetic Modeling Methods. CPT: Pharmacometrics & Systems Pharmacology. 2013;2(4):38. doi:10.1038/psp.2013.14.

21. Monolix version 2020R1. Antony, France: Lixosoft SAS; 2020. Available from: http://lixoft.com/products/monolix/.

22. Castro M, de Boer RJ. Testing structural identifiability by a simple scaling method. PLOS Computational Biology. 2020;16(11):1–15. doi:10.1371/journal.pcbi.1008248.

23. R Development Core Team. R: A Language and Environment for Statistical Computing; 2008. Available from: http://www.r-project.org.

24. Soetaert K, Petzoldt T, Setzer RW. Solving Differential Equations in R: Package deSolve. Journal of Statistical Software. 2010;33(9):1–25. doi:10.18637/jss.v033.i09.

25. Wickham H, Averick M, Bryan J, et al. Welcome to the Tidyverse. Journal of Open Source Software. 2019;4(43):1686. doi:10.21105/joss.01686.

26. Wilke CO. cowplot: Streamlined Plot Theme and Plot Annotations for ‘ggplot2’; 2020. Available from: https://CRAN.R-project.org/package=cowplot.

27. Pedersen TL. patchwork: The Composer of Plots; 2020. Available from: https://CRAN.R-project.org/package=patchwork.

28. Callaway D, Perelson A. HIV-1 infection and low steady state viral loads. Bull Math Biol. 2002;64(1):29–64. doi:10.1006/bulm.2001.0266.

29. Kuhn L, Paximadis M, Da Costa Dias B, et al. Predictors of cell-associated HIV-1 DNA over one year in very early treated infants. Clinical Infectious Diseases. 2021;ciab586. doi:10.1093/cid/ciab586.

30. Perelson AS, Neumann AU, Markowitz M, et al. HIV-1 dynamics in vivo: viron clearance rate, infected cell life-span, and viral generation time. Science. 1996;271(5255):1582–1586. doi:10.1126/science.271.5255.1582.

31. Luzuriaga K, Wu H, McManus M, et al. Dynamics of human immunodeficiency virus type 1 replication in vertically infected infants. J Virol. 1999;73(1):362–367.

32. Chun TW, Carruth L, Finzi D, et al. Quantification of latent tissue reservoirs and total body viral load in HIV-1 infection. Nature. 1997;387(6629):183–188.

33. Mehandru S, Poles MA, Tenner-Racz K, et al. Primary HIV-1 Infection Is Associated with Preferential Depletion of CD4+ T Lymphocytes from Effector Sites in the Gastrointestinal Tract. Journal of Experimental Medicine. 2004;200(6):761–770.

34. Pan X, Baldauf HM, Keppler OT, et al. Restrictions to HIV-1 replication in resting CD4+ T lymphocytes. Cell Res. 2013;23(7):876–85. doi:10.1038/cr.2013.74.

35. Zack JA, Kim SG, Vatakis DN. HIV restriction in quiescent CD4+ T cells. Retrovirology. 2013;10:37. doi:10.1186/1742-4690-10-37.

## References

1. Castro M, de Boer RJ. Testing structural identifiability by a simple scaling method. PLOS Computational Biology. 2020;16(11):1–15. doi:10.1371/journal.pcbi.1008248.

2. Prague M, Gerold JM, Balelli I, et al. Viral rebound kinetics following single and combination immunotherapy for HIV/SIV. bioRxiv. 2019;doi:10.1101/700401.

3. Monolix version 2020R1. Antony, France: Lixosoft SAS; 2020. Available from: http://lixoft.com/products/monolix/.

4. Mould D, Upton R. Basic Concepts in Population Modeling, Simulation, and Model-Based Drug Development – Part 2: Introduction to Pharmacokinetic Modeling Methods. CPT: Pharmacometrics & Systems Pharmacology. 2013;2(4):38. doi:10.1038/psp.2013.14.

